# A novel alpha-synuclein K58N missense variant in a patient with Parkinson’s disease

**DOI:** 10.1101/2025.02.07.25321793

**Authors:** Mohammed Al-Azzani, Sandrina Weber, Nagendran Ramalingam, Maria Ramón, Liana Shvachiy, Gonçalo Mestre, Michael Zech, Kevin Sicking, Alain Ibáñez de Opakua, Vidyashree Jayanthi, Leslie Amaral, Aishwarya Agarwal, Aswathy Chandran, Susana R. Chaves, Juliane Winkelmann, Claudia Trenkwalder, Maike Schwager, Silke Pauli, Ulf Dettmer, Claudio O. Fernández, Janin Lautenschläger, Markus Zweckstetter, Ruben Fernandez Busnadiego, Brit Mollenhauer, Tiago Fleming Outeiro

## Abstract

Mutations and multiplications in the SNCA *gene*, encoding alpha-synuclein (aSyn), are associated with familial forms of Parkinson’s disease (PD). We report the identification of a novel *SNCA* missense mutation (NM_000345.4, cDNA 174G>C; protein K58N) in a PD patient using whole exome sequencing, and describe comprehensive molecular and cellular analysss of the effects of this novel mutation. The patient exhibited typical sporadic PD with early onset and a benign disease course. Biophysical studies revealed that the K58N substitution causes local structural effects, disrupts binding to membranes, and enhances aSyn in vitro aggregation. K58N aSyn produces fewer inclusions per cell, and fails to undergo condensate formation. The mutation increases the cytoplasmic distribution of the protein, and has minimal effect on the dynamic reversibility of serine-129 phosphorylation. In total, the identification of this novel mutation advances our understanding of aSyn biology and pathobiology.

## Introduction

Parkinson’s disease (PD) is an age-associated neurodegenerative disorder that classically manifest clinically by a triad of cardinal motor symptoms of tremor, rigidity, and bradykinesia, as well as non-motor symptoms like depression, autonomic dysfunction and dementia (*1*, *2*). Neuropathologically, PD is marked by the progressive degeneration of dopaminergic neurons in in the substantia nigra and the presence of Lewy bodies (LBs), which are intraneuronal inclusions predominantly composed of aggregated alpha-synuclein (aSyn) (*3–6*). In addition to PD, misfolding and aggregation of aSyn into protein inclusions is present in other neurodegenerative disorders grouped together as synucleinopathies, such as dementia with Lewy bodies (DLB) and multiple system atrophy (MSA). Importantly, increasing evidence points to different structural organization of aSyn fibrils within inclusions that varies according to the synucleinopathy (*7–9*).

aSyn, an intrinsically disordered protein, has been a significant focus of neuroscience research not only due to its central role in PD and related synucleinopathies, but also because of its abundance and function in the nervous system. aSyn is primarily concentrated in the presynaptic terminals of neurons, but is also present in other cellular compartments including the nucleus, and is thought to interact with membranes adopting helical conformations and tetrameric structures (*10*, *11*). Nevertheless, the precise physiological functions of aSyn are still poorly understood, especially in what concerns its function in non-neuronal cells.

In pathological conditions, aSyn is thought to misfold and accumulate, forming pathological aggregates that disrupt cellular homeostasis, in a process that may ultimately cause neuronal cell death. Dopaminergic neurons in the substantia nigra seem particularly vulnerable to the toxic damage of pathological aSyn aggregates, although the underlying mechanisms remain unclear. While aSyn aggregation has been typically considered the driver of neurodegeneration, other theories suggest that the loss of functional monomeric aSyn due to aggregation, also known as synucleinopenia, may lead to disease progression (*12*, *13*). In reality, a combination of proteinopathy and proteinopenia are likely to contribute to disease.

Missense mutations in *SNCA*, the gene that encodes aSyn, are linked to familial forms of PD, suggesting that determining the molecular effects of such mutations may prove important for our understanding of the pathological underpinnings of the disease. In fact, since the discovery of the first *SNCA* mutation encoding for the A53T substitution (*3*), the field of PD genetics has enabled the identification of several other *SNCA* mutations as well as mutations in several other genes in PD (*14*, *15*). Several variants of aSyn have been linked to familial forms of PD and include, for example, G14R, V15A, A30G, A30P, E46K, H50Q, G51D, A53T, A53E, A53V, T72M, highlighting the importance of aSyn in PD (*3*, *6*, *16–24*). However, mutations in the *SNCA* gene are extremely rare and account for only 0.1-0.2% of cases (*14*). In addition, duplications or triplications of the chromosomal region containing the *SNCA* gene are also associated with familial forms of PD (*25*). These multiplications are consistent with a dose-dependent effect, as *SNCA* triplications cause a fully penetrant and severe phenotype with early onset and common non-motor symptoms like depression, psychosis and cognitive decline (*26*, *27*), while duplications on the other hand, are associated with a more variable clinical presentation, and more often present with Dementia with Lewy bodies rather than PD (*26*). Missense mutations are generally rarer than duplications (*15*, *28*), and are associated with a broad clinical spectrum that ranges from presentations resembling idiopathic PD, to more complex phenotypes, with atypical signs (*29*), resembling atypical PD or other neurodegenerative disorders. Consistently, our group has recently described the novel variant G14R, that is associated with an atypical, rapidly progressive phenotype and widespread neuronal loss in combination with frontotemporal lobar degeneration-associated aSyn pathology (*24*). To date, only a limited number of missense variants have been reported, including variants with unresolved pathogenic relevance and variants that have yet to undergo independent replication (*21*, *30*, *31*). In addition to the clinical heterogeneity, the identification of *SNCA* missense variants is hampered by the fact that some variants demonstrate incomplete penetrance (*16*, *32*), resulting in negative family histories. Consequently, validating known variants and identifying novel ones are crucial for improving diagnostic accuracy and for informing genetic counseling. Furthermore, *SNCA* missense variants are of tremendous interest for researchers because of their potential to expand our understanding of the molecular mechanisms underlying aSyn aggregation as well as pathological and physiological properties, including phosphorylation at S129 (pS129) (*33*, *34*). For instance, different missense variants have been associated with increased or decreased pathologic properties like aggregation and phosphorylation status (*24*, *35–37*). Thus, investigating these variants offers an invaluable opportunity to advance our knowledge of the physiological and pathophysiological roles of aSyn.

In this study, we identified a novel heterozygous missense mutation in a case of familial PD and provide a detailed description of the clinical phenotype and molecular effects.

## Results

### Clinical presentation

The index patient is a male Caucasian that presented initial motor symptoms approximately in his late 30s, with left-sided rest tremor, reduced arm swing and mild bradykinesia on the left side. He was diagnosed with early-onset PD (EOPD) in his early 40s. A dopamine transporter Imaging showed asymmetrical decrease consistent with PD, and no signs indicative of atypical PD or other neurologic disorders were found on clinical examination. After a good initial response to dopamine agonists and dopamine, the patient developed motor-fluctuations with end-of-dose wearing-off, trunk dyskinesias in the on-phase and freezing-of-gait. Marked dystonia was present in the left arm and leg. 13 years after PD diagnosis, he was evaluated for deep brain stimulation (DBS). At that time, non-motor symptoms included excessive daytime sleepiness, fatigue and depression. Comprehensive neuropsychologic assessment found no signs of cognitive dysfunction and a repeated MRI showed no pathologic findings or signs consistent with atypical PD. Presurgical levodopa-response as evaluated by levodopa-challenge test was highly effective, with an improvement of UPDRS-III of 78%. The patient received bilateral DBS of the subthalamic nucleus that reduced the severity of motor fluctuations, freezing, and tremor. Post-surgical follow-up 3 years after DBS implantation showed sustained good effects on motor-symptoms, overall good cognition with MMSE 30/30 but re-emergence of moderate motor-fluctuations. The family history was positive, and both a parent and a grandparent were affected by PD. Both relatives were already deceased but were reported to have had no atypical symptoms and no dementia.

### Whole-exome sequencing identifies aSynp.K58N point mutation

The patient was selected for genetic testing by whole-exome sequencing (WES) due to the early onset of PD and the positive family history. WES identified a novel heterozygous missense variant in *SNCA* (NM_000345.4, cDNA 174G>C; protein K58N) (Fig. S1 A and B). The variant was not annotated in the genomic databases gnomAD and the PD Variant Browser. It causes a missense substitution in a phylogenetically highly conserved region (Fig. S1 C) and is predicted as deleterious by several in-silico prediction algorithms (CADD:31, PolyPhen-2: 0.994, PrimateAI: 0.8337, REVEL: 0.653).

### K58N mutation causes a reduction in α-helical content, impairing its interaction with liposomes

In order to study the impact of K58N mutation on the structural and membrane-binding properties characteristics of aSyn, we employed NMR spectroscopy and circular dichroism (CD) using recombinantly prepared monomeric WT and K58N aSyn. The two-dimensional NMR ^1^H/^15^N- correlation spectra for both proteins revealed minimal signal spread as expected for IDPs (Fig. 1 A). Spectral comparison showed that the majority of the cross peaks for both variants aligned together except for those linked to residues located around the mutation site (Fig. 1 B-D). The disturbances in the NMR signal shifts and intensity were restricted to the vicinity of the K58N mutation as shown in (Fig. 1 B-D).

**Fig. 1.**
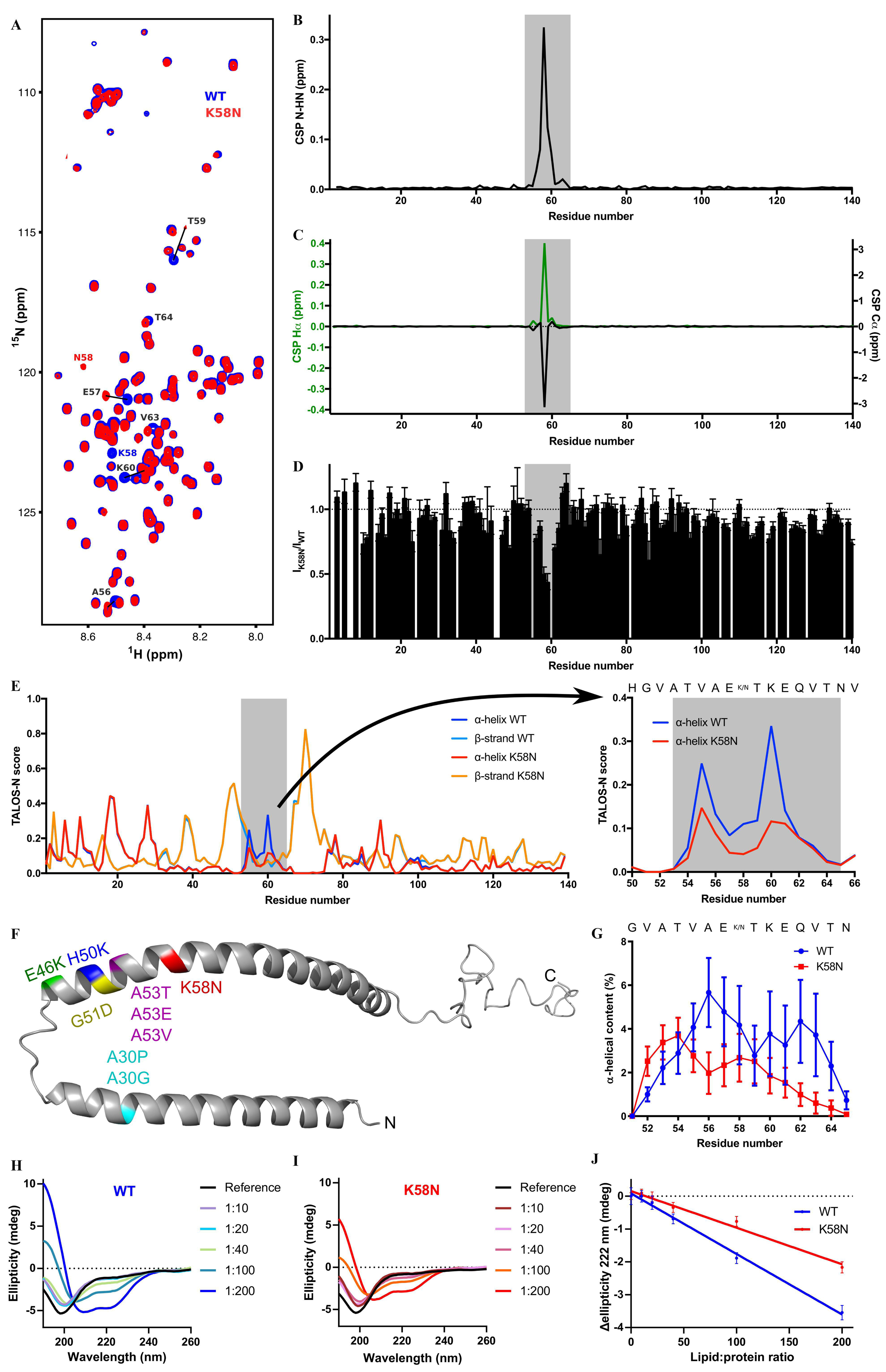
K58N mutation decreases the α-helical content affecting liposome binding. a, ^1^H,^15^N HSQC of αSyn WT (blue) and K58N (red). Evident CSPs are labeled. b, Combined HN/N CSPs generated by the K58N mutation over the αSyn sequence. Affected region is highlighted in grey. c, Hα (green) and Cα (black) CSPs. d, Normalized ^1^H,^15^N HSQC intensity ratios. Error bars are calculated from signal-to-noise ratios of individual resonances. e, Secondary structure calculations from NMR chemical shifts for αSyn WT (α-helix, blue; β-strand, light blue) and K58N (α-helix, red; β-strand, orange). On the right a zoom of the affected region with the α- helical content is plotted. f, Micelle-bound αSyn structure (PDB ID: 1XQ8) with the positions of the different pathological mutations highlighted. g, Residue specific α-helical content over 100 ns MD simulations. Error bars indicate the standard error from 40 analyzed peptides. h-i, CD experiments of WT (h) and K58N (i) αSyn at different protein:lipid ratios. j, Ellipticity change at 222 nm upon increasing concentrations of lipids for WT (blue) and K58N (red). More negative values indicate a bigger increase of α-helical structure.

To gain insight into how this could affect the local conformation of K58N aSyn, we calculated the secondary structure from NMR chemical shifts around the mutation area. Interestingly, we observed a reduction in the amount of α-helical content with the K58N mutation in that region (Fig. 1 E). Furthermore, MD simulations of the aSyn peptide G51-N65 either containing the WT lysine or an asparagine at position 58 reproduced the difference observed in the experimental NMR data of the full-length protein showing a decrease in α-helix content for the mutant, albeit with different total values (Fig. 1 F). The findings from both experimental and simulation demonstrate that K58N mutation impact the helical propensity surrounding the mutation site (Fig. 1 E and G).

aSyn is known to form α-helical structures upon binding to membranes. Therefore, we investigated the effect of K58N mutation on its interactions with membranes by incubating both WT and K58N aSyn with increasing amounts of liposomes and utilizing CD, a well-established technique for monitoring the transition of aSyn into membrane-bound α-helical conformations (*38–40*). The exposure to liposomes resulted in a shift from random coil into α-helical structure for both variants (Fig. 1 H and I). However, analysis of the change in the ellipticity at 222 nm revealed that K58N mutation exhibited a lower tendency to adopt α-helical conformations when exposed to liposomes reflecting a reduction of the binding to liposomes (Fig. 1 J). In summary, the NMR data shows that the α**-**helical content is reduced by the mutation in the monomeric state, which may explain the decreased liposome binding, as the liposome-bound aSyn tends to form alpha-helix in that region.

### Effect of the K58N mutation on aSyn aggregation in vitro

A central pathological hallmark in PD and other synucleinopathies is the accumulation of aSyn into insoluble aggregates. Therefore, we investigated the aggregation propensity of the K58N variant in comparison to WT aSyn. To check this, we initially assessed the aggregation propensity of K58N variant employing various computational prediction algorithms (*41–43*). These algorithms showed increase in β-strand content, and a higher tendency in the aggregation properties for K58N aSyn (Fig. S2). Experimentally, recombinant WT and K58N aSyn were incubated under shaking conditions to monitor the fibrillization kinetics of both proteins, following an in vitro ThT-based aggregation assay protocol. Interestingly, the K58N showed a higher ThT aggregation profile compared with WT aSyn (Fig. 2 A-B and D). Furthermore, the K58N variant demonstrated different aggregation kinetics, with a shorter t_1/2_ suggesting a faster rate of fibril formation (Fig. 2 C). These findings prompted us to perform detailed cryo-EM analysis of the morphological, organizational, and structural features of fibrils prepared by both variants. As previously reported (*24*), WT a-Syn fibrils adopted two similarly populated conformations, consisting of one (1PF) or two protofilaments (2PF) respectively (Fig. 3A, C; Fig. S3). 3D reconstruction was only successful for 2PF fibrils, resulting in a structure with 2.7 Å global resolution (Fig. 3 E; Fig. S3; Fig. S4) that enabled building of an atomic model. This model displays a double-arrow fold previously observed in other studies, such as “polymorph 2A” reported by (*44*). Similar to that structure, our model features a salt bridge between lysine 45 (K45) and glutamic acid 57 (E57) that stabilizes the inter-protofilament interface (Fig. 3G, H). The protofilament fold itself is nearly identical to those described as “polymorph L2A and L2B” by (*45*) and “protofilament fold B” by (*46*). However, in these cases, the inter-protofilament interactions differ.

**Fig. 2.**
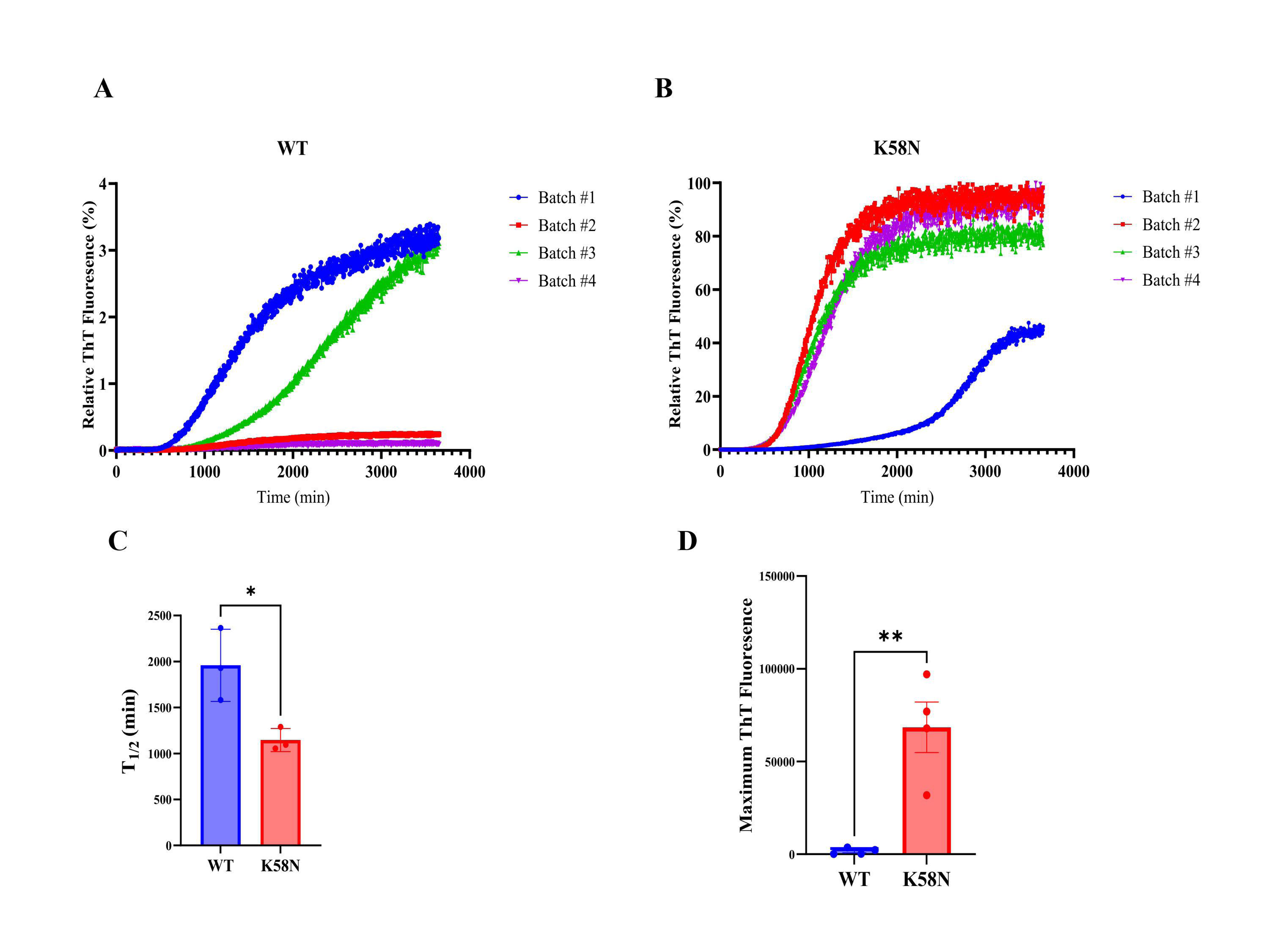
In vitro ThT-based aggregation assays for WT and K58N aSyn. (A) and (B) Aggregation kinetic profiles of WT and K58N aSyn. Data was normalized to the maximum fluorescence value of each run. (C) Comparison of the half-time in min to for aggregation kinetics. (D) Comparison of maximum ThT fluorescence values of WT and K58N. Error bars represent mean±SD.

**Fig. 3.**
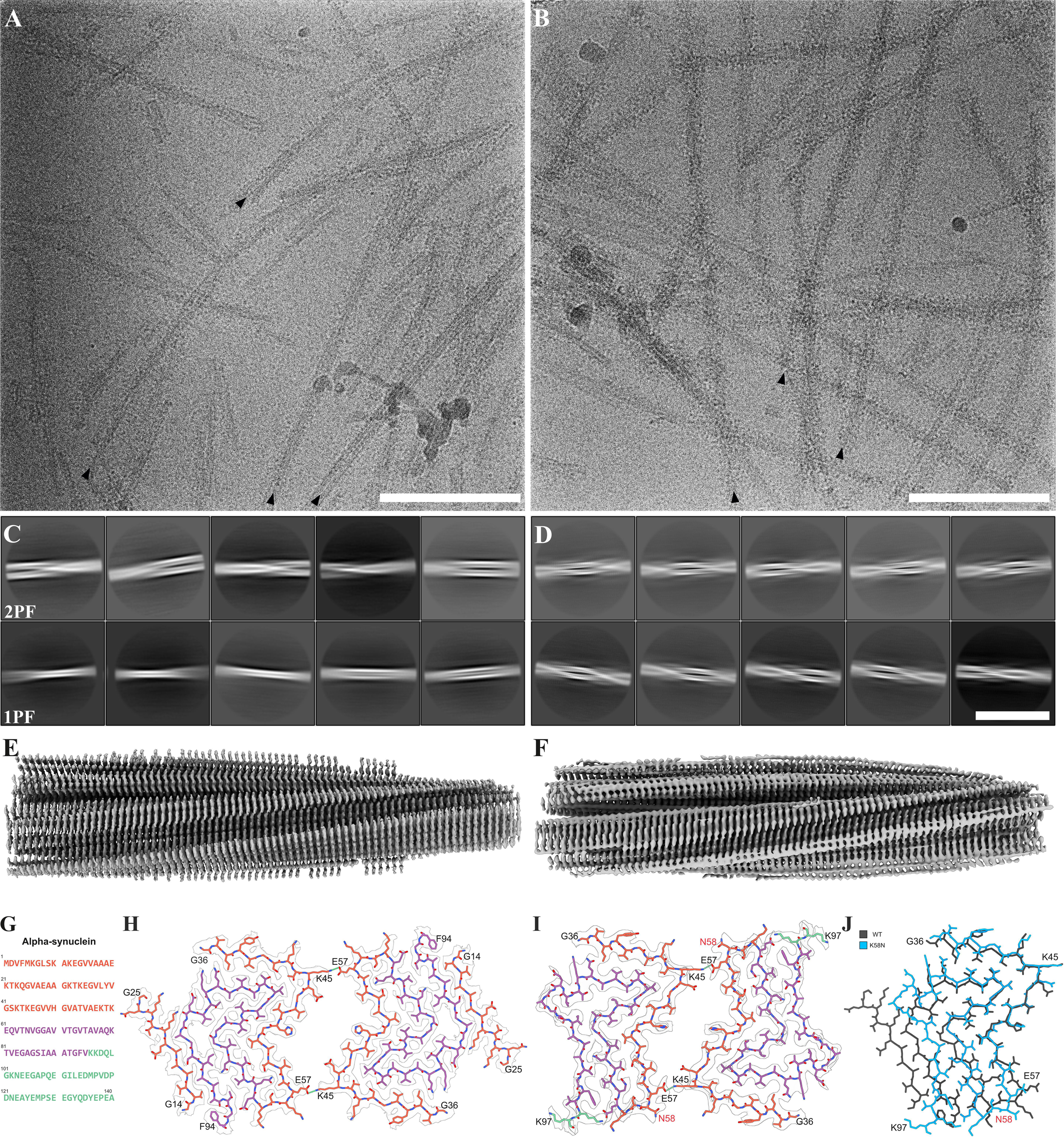
Characteristics of WT and K58N aSyn filaments. A) TEM micrograph of WT aSyn amyloid filaments. Black arrows mark selected filament ends. Scale bar: 100 nm. B) TEM micrograph of K58N aSyn amyloid filaments. Black arrows indicate filament ends of exemplary filaments. Scale bar: 100 nm. C) 2D class averages (706 Å box size) of twisting WT aSyn filaments, illustrating the interaction between either two protofilaments (2PF) or a single protofilament (1PF). Scale bar: 50 nm. D) 2D class averages (706 Å box size) reveal twisting K58N aSyn filaments, again characterized by two protofilaments interacting with each other. Scale bar: 50 nm. E) Overview of the electron density map of WT aSyn filaments. F) Overview of the electron density map of K58N aSyn filaments. G) Amino acid sequence of human aSyn with distinct regions color-coded (N-Terminus in orange, Non-amyloid component in purple, and C- Terminus in green). H) The electron density map together with the atomic model of WT aSyn amyloid filaments featuring a single beta-sheet layer formed by two interacting protofilaments. The protofilament interface is stabilized by a K45-E57 salt bridge. I) The electron density map together with the atomic model of K58N aSyn amyloid filaments. The protofilament interface is again stabilized by the K45-E57 salt bridge. The mutated residue is indicated in red. J) Superposition of the WT atomic model (grey) and the K58N mutant (blue). While the double-arrow structure is similar, notable differences in the backbone highlight the impact of the K58N mutation (residue marked in red) on the overall aSyn filament structure.

In contrast, K58N adopted almost exclusively 2PF conformations (Fig. 3B, D; Fig. S5). An atomic model based on the 3D reconstruction of these fibrils at 3.7 Å global resolution revealed a similar protofilament fold as in WT fibrils (Fig. 3F, I, J, Fig. S4). However, K58N fibrils present a higher twist (−1.29 degrees vs −0.81 degrees in WT fibrils), and a slightly shifted protofilament interface, possibly due to the proximity of the mutation site (K58) to the interface stabilizing residue (E57). Additionally, no density for residues 14-25 was visible at the periphery of mutant fibrils (Fig. 3G-J). Altogether, these data indicate that the K58N mutation displaces the conformational equilibrium of a-Syn fibrils towards a 2PF structure reminiscent of that observed for WT.

### K58N mutation alters aSyn inclusion number and size in a cellular model

After studying the impact of K58N mutation on aggregation of aSyn in vitro, we next explored whether a similar aggregation tendency can be observed under cellular environment. To this end, we employed the SynT/Sph1 model, a well-recognized methodology that has been used regularly to study aSyn aggregation in cells (*47*). Human neuroglioma cells (H4) were first co-transfected with WT or K58N SynT variants and Sph1 followed by immunostaining to assess inclusion formation 48 hours post-transfection. Interestingly, cells expressing K58N SynT mutation exhibited a significant decrease in the number of aSyn inclusions in comparison to cells expressing WT aSyn (Fig. 4 A and B). The inclusions observed in K58N transfected cells were smaller in size (Fig. 4 C). Moreover, a significant increase in the percentage of cells without inclusions was observed in cells expressing K58N version (Fig. S6).

**Fig. 4.**
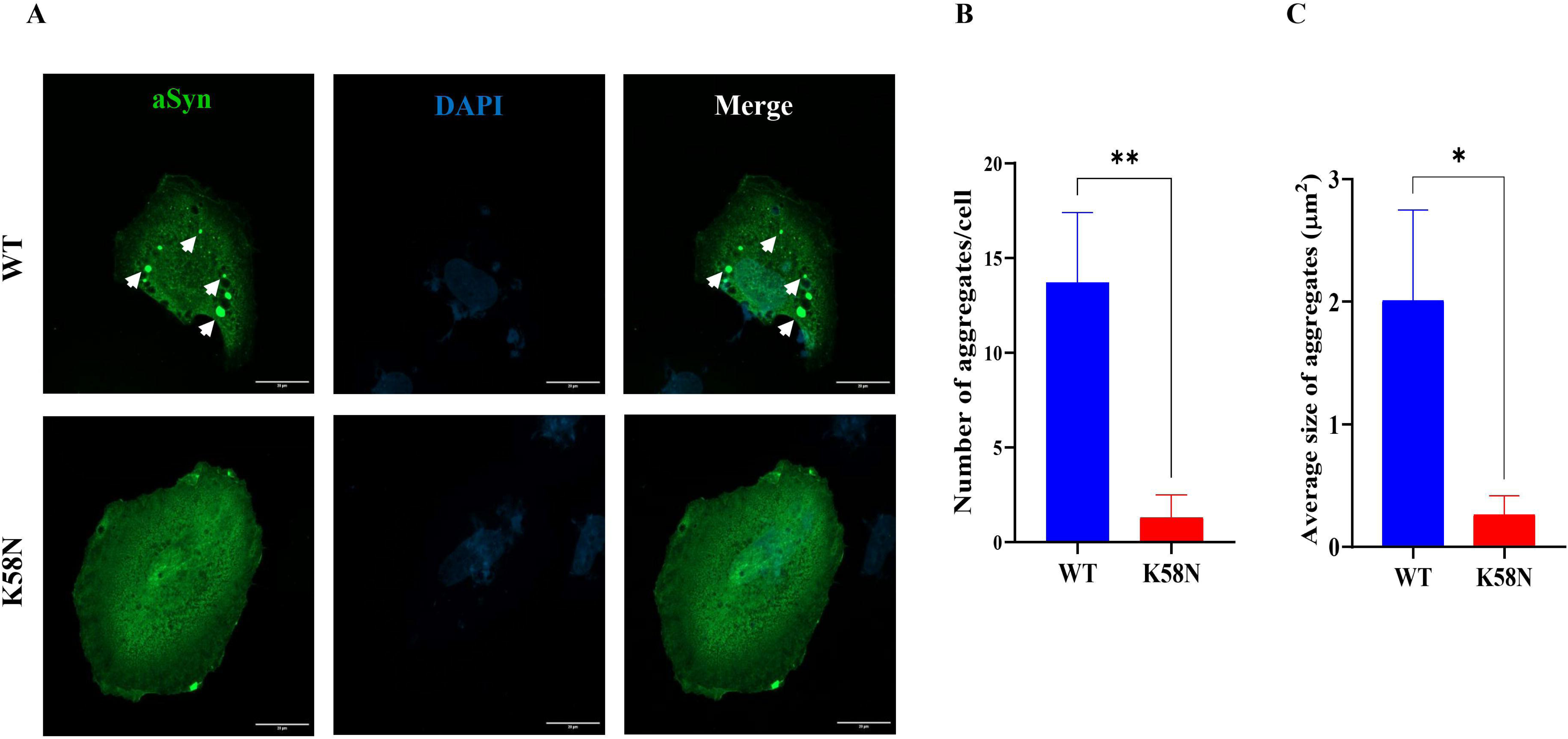
Effect of K58N mutation on inclusion formation in cells. The pattents of inclusions formation was enivtagated using SynT/Sph1 aggregation model, which is based on the co-expression of based on the co-expression of WT or K58N SynT variants and synphilin-1. (A) Immunohistochemistry images representing the inclusion formation in H4 cells expressing WT and G14R aSyn. Scale bar: 20 µm. (B) Quantification of the number of inclusions per cell and their area (C). For each experiment, 50 cells were counted at a 100x magnification. The data analysis was performed using a Student’s t-test, and presented as mean ± SD (*N=3*).

### K58N aSyn shows decreased phase separation

We further explored the impact of this novel mutation on aSyn phase separation properties, a critical factor that is thought to play a role in its functional and pathological behavior. We find that K58N aSyn undergoes only minimal droplet formation compared to aSyn wild-type (WT) when tested under otherwise same conditions (Fig. 5 A). This is recapitulated at a quantitative scale, testing aSynK58N phase separation using turbidity measurements in the presence of Ca^2+^ and varying PEG concentrations (Fig. 5 B).

**Fig. 5.**
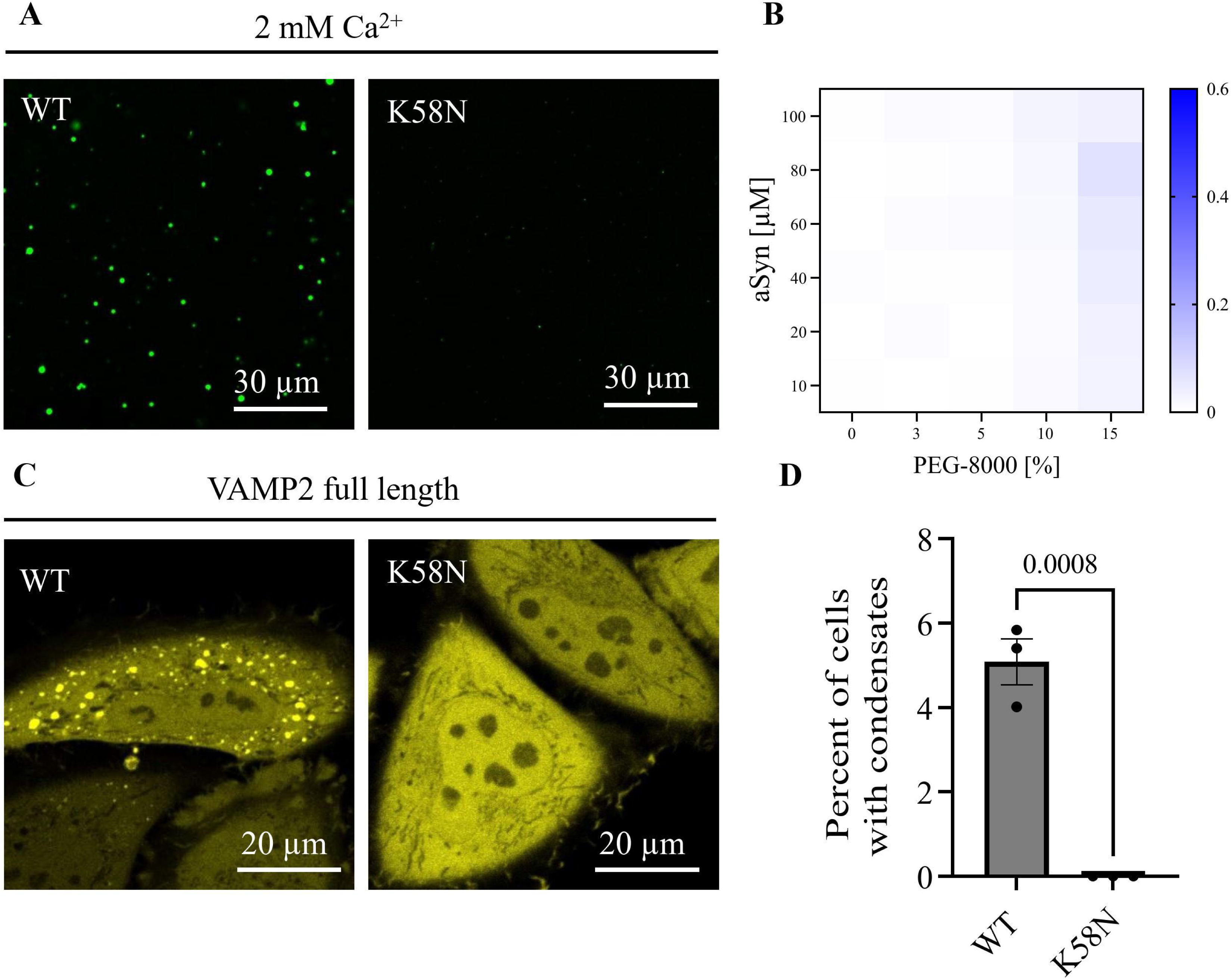
aSyn K58N shows decreased condensate formation in vitro and in cells. (A) aSyn phase separation in the presence of 2 mM Ca^2+^ and crowding with 15% PEG-8000, immediately after PEG addition for aSyn wildtype (WT) and the disease variant aSyn K58N. aSyn concentration used: 100 µM. (B) Heatmap for turbidity measurements of aSyn phase separation in the presence of 2 mM Ca^2+^. Data derived from 4 independent repeats. (C) Condensate formation of aSyn WT YFP and aSyn K58N YFP upon ectopic expression with VAMP2 in HeLa cells. aSyn K58N YFP does not undergo condensate formation in cells. (D) Quantification of condensate formation. Data derived from incuCyte screening, 16 images per well, 3 wells per biological repeat, 3 biological repeats. n indicates biological repeats. Data are represented as mean +/- SD. Unpaired two-tailed t-test.

We next expressed aSyn YFP with VAMP2 in HeLa cells, where VAMP2 induces aSyn phase separation as previously shown (*48*). Here, WT aSyn YFP shows condensate formation, however K58N aSyn YFP fails to form condensates exhibiting a homogenous cytosolic distribution (Fig. 5 C). Quantitative evaluation confirms the absence of condensate formation for the K58N aSyn variant (Fig. 5 D).

### Effects of the K58N mutation on aSyn S129 phosphorylation

There is increasing evidence that serine-129 phosphorylation (pS129) of aSyn is not only associated with pathology but also influences the physiological function of aSyn (*34*). Therefore, we investigated whether the K58N mutation alters the pS129 status. To address this, we employed lentiviral vectors to express either human WT or K58N variants in primary aSyn knockout (*SNCA*−/−) rat cortical cultures (Fig. 6 A). We first checked the levels of pS129 in the absence of neuronal stimulation. Compared to WT aSyn, K58N mutant exhibited a significant reduction in basal pS129 (Fig. 6 B). Recent studies have shown that familial PD-associated aSyn mutants with reduced membrane (M) localization is often associated with lower basal pS129 levels (*34*). Given the observed decrease in pS129 levels observed here for K58N mutant, we hypothesized that this decrease could be attributed to increased solubility or, in other words, decreased membrane association (M). In line with this, we observed that the K58N mutation was enriched in the cytosolic fraction (C) (GAPDH fraction, ∼ 62%) (Fig. 6 C and D), supporting the idea that reduced membrane association contributes to lower pS129 levels.

**Fig. 6.**
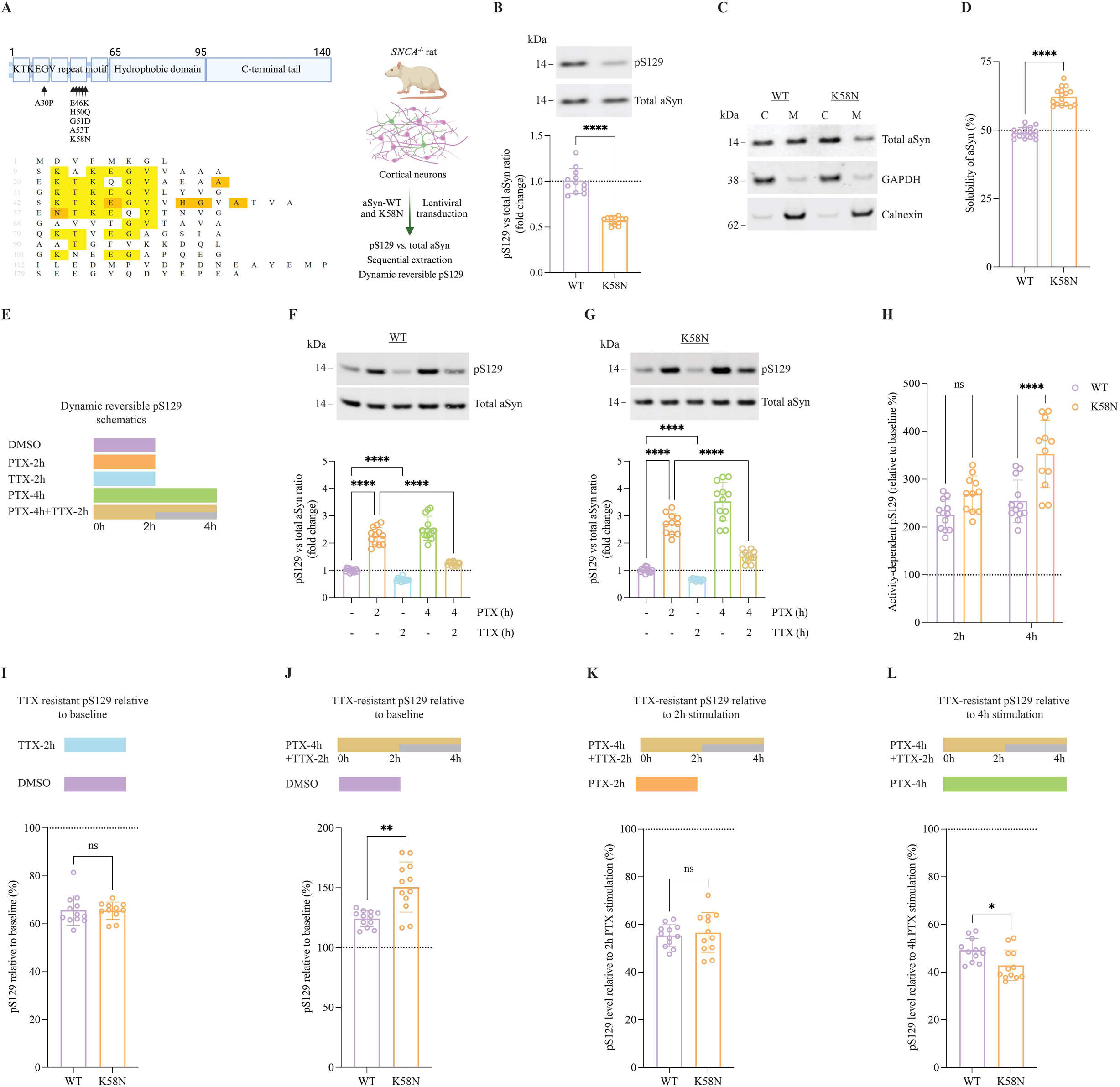
Dynamic activity-dependent pS129 of K58N and WT aSyn. (A) Schematic illustration of aSyn structure exhibiting the KTKEGV repeat sequence harboring most familial PD mutations, central hydrophobic domain, and C-terminal region. The sequence alignment of aSyn is displayed below showing the conserved KTKEGV residues in yellow and sites of familial PD mutation in orange. On the right, the experimental setup is summarized. (B) A representative western blot (WB) displaying the levels of total aSyn and pS129 from DIV17-21 rat *SNCA*^−/−^ rat cortical neurons expressing WT and G14R aSyn, with WB quantitative analysis presented below. (C) WB results of WT and G14R transduced rat *SNCA*^−/−^ cortical neurons that were subjected to on-plate sequential extraction to isolate the cytosolic (C) and membrane (M) fractions. MJFR1 antibody was used for total aSyn detection, while GAPDH and Calnexin were used as cytosolic and membrane fractions controls, respectively. (D) quantitative analysis of WT and K58N aSyn solubility from WB data in (C). (E) A summary of the experimental conditions to study pS129 dynamic reversibility, with more information are provided in the main text. (F-G) Neuronal activity-dependent reversible phosphorylation of S129 (as outlined in schematic E) was detected in DIV17-21 rat *SNCA−/−* cortical neurons expressing WT or G14R, respectively, following PTX stimulation (20 µM) and TTX inhibition (1 µM). WB for quantifying total aSyn and pS129 was employed and panels H-L are derived from F to G. (H) The percentage of increase in pS129, compared to baseline, for WT and G14R aSyn observed after 2h or 4h PTX stimulation. (I) Comparison of TTX-resistant pS129 levels in WT and G14R variants relative to baseline conditions (DMSO vehicle). (J) The proportion of irreversible pS129 relative to the basal (DMSO vehicle) condition. (K) The proportion of irreversible pS129 relative to 2 h PTX stimulation. (L) The proportion of irreversible pS129 relative to 4 h PTX stimulation. \*\*\*\**P* < 0·0001; \*\*\**P* < 0·001; \**P* < 0·05; ns, not significant. The error bar was mean± SD.

Phosphorylation of S129 also has been recently reported to be an activity dependent, dynamic physiological process. According to these studies, pS129 is elevated following neuronal stimulation and then returns to baseline once the stimulus dampens or when inhibited in a dynamic reversible process (*34*). Importantly, rodent neuron cultures expressing PD-associated A30P and E46K aSyn mutants showed impaired dynamic reversibility of pS129 (*49*). To assess the effect of the K58N PD-associated mutation on the dynamic reversibility of pS129, we exposed WT or K58N aSyn transduced rat *SNCA*−/− cortical neurons to neuronal stimulation, inhibition, or a combination of stimulation followed by inhibition (Fig. 6 E). At DIV 17-21, cortical cultures were stimulated with picrotoxin (PTX), a GABA_A_ receptor antagonist, to stimulate neuronal activity and inhibited by the sodium channel blocker, tetrodotoxin (TTX). Consistent with our previous observations, the exposure of neurons to PTX for 2h and 4h significantly increased pS129 levels in WT neurons, while neuronal inhibition through TTX decreased the basal levels by approximately 30%. The TTX inhibition 2h post-PTX stimulation effectively reversed activity-dependent pS129 elevation (Fig. 6 F). K58N neurons followed a comparable pattern of activity-dependent pS129 response (Fig. 6 G). Interestingly, however, PTX treatment for 4h induced a more pronounced increase in pS129 in K58N aSyn neurons compared to that of WT, relative to basal levels (Fig. 6 H). Then, we compared the 4h PTX stimulation combined with TTX inhibition introduced midway to 2h or 4h PTX stimulation in order to evaluate the dynamic reversibility of pS129. The percentage of irreversible pS129 levels in PTX/TTX-treated neurons was comparable between K58N and WT aSyn relative to PTX stimulation for 2h (Fig. 6 K). However, after 4h of PTX stimulation, a lower proportion of irreversible in K58N neurons compared to WT aSyn expressing neurons (Fig. 6 L). We assessed the sensitivity of K58N transduced neurons to TTX inhibition under unstimulated conditions and found no significant differences between K58N and WT neurons (Fig. 6 I). In conclusion, our findings indicate that activity-dependent pS129 is altered in the disease-associated K58N aSyn mutant, with a pronounced effect in response to prolonged neuronal activity.

### K58N is more cytosolic without changing cytotoxicity in yeast cells

To investigate how the K58N aSyn mutation affects cellular distribution and cytotoxicity, we used the well-established budding yeast model to associate subcellular localization with toxicity (*50*). In our results we observed that the K58N aSyn is predominantly localized to the cytoplasm in yeast cells, in contrast to WT aSyn, which tended to form cytoplasmic inclusions (Fig. 7 A). Despite the different subcellular localization, the K58N and WT aSyn presented identical toxicities (Fig. 7 B).

**Fig. 7.**
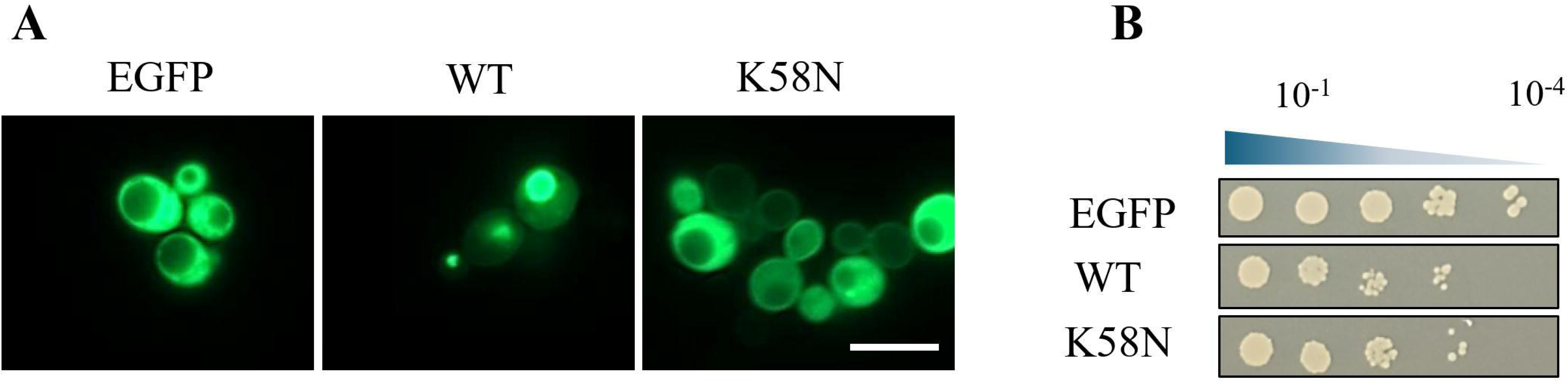
Effect of K58N mutation on yeast cells. S. cerevisiae cells harboring WT aSyn or K58N aSyn mutation were grown to mid-log phase. (A) aSyn localization was analyzed by fluorescence microscopy. (B) Cellular viability was evaluated by spotting assay, where cultures were spotted on SD-URA agar plates.

## Discussion

Here, we report the discovery of missense mutation K58N in the *SNCA* gene and provide a detailed description of the associated clinical phenotype and molecular effects of this novel mutation. The variant was identified through WES in a patient with EOPD and a positive family history, both of which raised the suspicion of an underlying genetic cause. Although both affected relatives were already deceased, and no DNA was available to confirm the mutation, the pathogenic relevance of K58N is supported by (1) the absence of K58N in genomic databases, (2) the disruption of a highly conserved amino acid residue, (3) the prediction of a pathogenic effect by different in-silico tools, and most importantly, (4) the results of the molecular characterization, that are discusses in detail further down. According to ACME (American College of Medical Genetics) guidelines (*51*), the variant is classified as *likely pathogenic*.

The clinical presentation and progression were similar to idiopathic PD, with a good and sustained levodopa-response and the lack of motor- and non-motor-symptoms indicating atypical PD. Both father and grandfather were also diagnosed with PD and even though both were already diseased and could not be examined, the medical history provided by the index patient was in accordance with idiopathic PD. Regarding the phenotypic spectrum of known *SNCA* missense mutations, manifestations that are indistinguishable from idiopathic PD as well as atypical and more complex syndromes have been described (*29*). While atypical symptoms are for example common in carriers of G51D (*18*, *52*), they are rarely described in carriers of A30P and A30G (*22*, *53*). Additionally, both A30P and A30G seem to lead to an overall more benign disease course compared to other missense mutations, which is similar to our identified mutation. However, cognitive decline was a common feature in both carriers of A30P and A30G and could even precede the onset of motor-symptoms, which was not observed in our present case. Another feature that is often seen across all *SNCA* mutations (including multiplications) are motor-fluctuations, that are present in around tree thirds of cases (*26*). In the patient with K58N, the severity of motor-fluctuations eventually led to the implantation of bilateral nucleus subthalamicus DBS. The patient profited from this intervention, with a good overall effect on motor-symptoms and motor-fluctuations. Thus, our study provides valuable information about beneficial DBS in K58N *SNCA*, even though the long-term effect has yet to be assessed, as motor-fluctuations seemed to re-emerge 4 years after DBS. A growing body of research suggest that the outcome of DBS is influenced by the presence of different genetic mutations (*54*). So far, data on DBS in *SNCA* missense variants is rare and limited to a few cases, with variable results (*22*, *55–58*). Whilst all cases had good short-term benefits from subthalamic nucleus DBS, especially on motor-fluctuations, two unrelated patients with A53E experienced rapid progression of motor- and non-motor symptoms after surgery (*55*, *57*). Re-emergence of motor-fluctuations after DBS was also reported in two patients with A30G (after 3 years) (*22*) and E46K (unknown interval) (*58*), respectively, but both cases had an overall less aggressive progression than the A53E cases. Additional data is needed to elucidate the relationship between *SNCA* mutations and DBS outcome, as this has important implications for patient counseling and indication for DBS. Most aSyn PD-associated mutations identified thus far occur in the N-terminal domain of aSyn. This region plays a crucial role in the interaction of a Syn with lipids and biological membranes. Binding to membranes is mediated through the repeated KTKEGV motif that is essential for the formation of an amphipathic α-helical structure needed for the interactions with membranes, which ultimately affect several structural and functional aspects of aSyn in cells. For example, aSyn exists, under normal conditions, in a dynamic equilibrium between its cytosolic soluble disordered form and helically structured membrane-bound conformations (*59*, *60*). Interestingly, most PD known mutations are present in the N-terminal domain region of aSyn leading to conformational alterations affecting its helical propensity, and therefore its preferential binding to membranes versus its solubility in the cytosol, und ultimately functional and pathological properties (*47*, *61*). The K58N mutation is also located in the N-terminal region and, more importantly, within the KTKEGV sequence. Lysines (K) are charged residues that play important role in the formation of amphipathic α-helices by establishing electrostatics interactions the with negatively charged heads of the lipids in the membranes. The replacements of the charged residue (K) within the KTKEGV sequence with uncharged residue asparagine (N) is expected to disrupt α-helix formation and electrostatic interactions with membranes (*62*). Furthermore, asparagine residues tend to occur in bends, turns and random coils. Consistently, our NMR data showed a local effect for the K58N mutation that was found mainly around the mutation site, which decrease the α-helical content of residues A53-N65. Simulation data were complementary to NMR findings, further supporting the idea that the K58N mutation disrupts the helical propensity of aSyn around the mutation site. The mutation-induced alterations in conformational dynamics are expected therefore to influence several functional properties for aSyn including its interactions with membranes. Despite this, the extent to which each PD-associated mutation alters membrane binding can differ from one mutation to another mutation, and can be influenced by other factors including the lipid composition and the curvature of those membranes. Among PD mutations, A30P is known to exhibit defective binding to membranes due to its decreased helical propensity (*63*, *64*). In the current study, we observed that the α-helical propensity for K58N is reduced in the presence of liposomes when compared to WT aSyn, consistent with a lower affinity of K58N to lipids.

A central pathological hallmark of various synucleinopathies is misfolding and aggregation of aSyn into insoluble and fibrillar protein deposits. Furthermore, different fibrils strains have been observed in certain synucleinopathies. Therefore, we investigated the impact of the K58N mutation on aSyn assembly. Importantly, the mutation occurs in a residue that is very close the central hydrophobic domain which plays a crucial role in the aggregation of aSyn. Furthermore, the local effects induced by this mutation were not limited to the immediate proximity of the mutation site but also residues further away, including some in the beginning of the central hydrophobic domain. According to different aggregation prediction algorithms, it is expected that K58N mutant aSyn exhibits higher aggregation propensity. Interestingly, this is consistent with our experimental in vitro aggregation assays that showed increased ThT binding and different aggregation kinetics when compared to WT aSyn. It is possible that the positively charged K at 58 position protects against aggregation, and that this effect is lost once it is mutated to a neutral N residue. This is consistent with data from studies of the H50Q mutation, as both substitutions have similar charge effects. Histidine at position 50 carries a partial positive charge at neutral pH, and it is substituted by a neutral Q leading to increased aggregation tendency in vitro (*65*, *66*).

The observed changes may also reflect differences in the structural characteristics of the fibrils between WT and K58N aSyn. The findings from cryo-EM data revealed that the K58N mutation preferentially results in 2PF fibrils with a similar protofilament fold than WT fibrils, suggesting that the mutation stabilizes the WT inter-protofilament interface. One possibility is that K58 modulates the ability of the adjacent interface-stabilizing residue (E57) to interact in cis or trans. In WT, K58 may form a salt bridge in cis with E57, which could destabilize the inter-protofilament interface and thereby favour 1PF structures. In contrast, the K58N mutation may prevent this interaction due to the altered charge properties, thereby promoting trans interactions with K45 of an adjacent protofilament and thus favoring in 2PF fibrils. Interestingly, the effects of the K58N mutation are opposite to those we recently reported for the G14R mutation (*24*), which strongly favors 1PF forms. Altogether, these results indicate that the main effect of these mutations in fibril architecture is the modulation of the WT conformational landscape, favoring either 1PF (G14R) or 2PF (K58N) forms.

In cells, K58N aSyn formed fewer inclusions when compared to WT aSyn and, overall these inclusions were smaller. These data are not surprising as some aSyn mutations previously studied showed, for example, attenuated aSyn aggregation in vitro while exhibiting increased formation of cellular inclusions (*24*, *47*). The findings using the yeast model revealed a more diffuse cytoplasmic distribution, further demonstrating the importance of employing diverse experimental models when evaluating the effects of mutations in the behaviour of aSyn, as this will likely uncover different properties of the protein.

The process of aSyn phase separation, leading to the formation of condensates (*67*, *68*), is considered dynamic under physiological conditions, but it has been shown that aSyn phase separation, under certain pathological states, can result in the formation of precursors or seeds that transform into solid, insoluble pathological aggregates (*69–71*). More recently, it has been shown that aSyn phase separation and condensate formation is regulated by VAMP2 (*48*, *72*), and that this regulatory mechanism may rely on the binding of aSyn to lipid membranes (*48*). In this assay, the A30P aSyn mutant, which is typically known for its defective lipid-binding properties (*62*, *73–75*) and increased cytoplasmic localization (*50*, *76*), failed to form condensates in cells. Interestingly, several assays conducted here revealed that the K58N mutant exhibits disrupted binding to membranes and increased cytosolic distribution, similarly to what was observed with A30P.

The phosphorylation of aSyn at serine 129 (pS129), previously recognized as pathological hallmark (*33*), is increasingly considered to also play a physiological role (*34*, *77*). This phosphorylation is a dynamic and reversible neuronal activity-dependent process that increases when neurons are stimulated, and is reversed when neuronal activity is reduced (*34*). In our study, we observed similar patterns of activity-dependent pS129 for both WT and K58N aSyn, in agreement with previous findings for some PD-associated mutations (*24*, *49*). These findings are consistent with different pathological effects by different mutations, and is consistent with a more benign disease course in the affected patient. Nevertheless, our study points to the importance of assessing the dynamic reversibility of S129 phosphorylation, as this sheds light into the molecular effects of aSyn mutations.

In conclusion, we report the identification of a novel heterozygous *SNCA* mutation, encoding for K58N aSyn, that is characterized clinically by causing early onset, and a benign course of PD that showed good response to DBS. The mutant protein was found to exhibit unique molecular and functional properties, such as reduced helical propensity, impaired lipid binding, increased solubility, and reduced condensation and inclusion formation in cells. Ultimately, our findings advance our understanding of the biology and pathobiology of aSyn, and provide new insight into the molecular mechanisms underlying PD and other synucleinopathies.

## Materials and methods

The patient was evaluated by neurologists specialized in movement disorders at the tertiary movement disorder center Paracelsus Elena-Klinik, Kassel, Germany. Repeated general and neurologic examinations included assessment of motor-symptoms by UPDRS, cognition by MMST, standardized levodopa challenge as described previously. Written informed consent was given by the patient to participate in a genetic study in cooperation with the Institute of Human Genetics, Technical University Munich, Germany. The study was approved by local ethic committees. Additional consent to confirm the genetic finding at the Institute of Human Genetics, University Clinic Göttingen, Germany was provided. Deep brain surgery (DBS) was performed at the Clinic of Neurosurgery, University Medical Center Göttingen, Germany.

### Genetic testing

Whole-exome sequencing (WES) was performed at the Institute of Human Genetics at the Technical University of Munich, Germany. Genomic DNA was extracted from peripheral blood according to standard protocols. In-solution enrichment of targeted regions was accomplished using the Sure Select Human All Exon Kit (Agilent 60mb V6) and followed by paired-end sequencing of 100 base pare (bp) long reads with the Illumina NovaSeq6000 system (Illumina, San Diego, California). Average exome coverage was 102x, with 97% of the target regions being covered at least 20x. The whole *SNCA* region was covered >20x (average depth 80x). The mutation was confirmed in an independent blood sample by Sanger sequencing by the Human genetics Department at the University Medical Center Göttingen.

### Expression and purification of recombinant WT and K58N aSyn

The K58N mutation was incorporated into the bacterial plasmid encoding aSyn using through site-directed mutagenesis (QuikChange II, Agilent), and Sanger sequencing was used to validate the presence of the mutation. The production of recombinant proteins was performed in BL21(DE3) *E. coli* cells that were transformed with pET21A vectors encoding either WT or G14R aSyn, following established protocols (*78*). After extracting proteins from bacterial pellets, they were purified through sequential anion-exchange and size-exclusion chromatography (SEC) for use in aggregation assays, nuclear magnetic resonance (NMR) analysis, and lipid binding experiments. The purified proteins were concentrated in PBS buffer (pH 7.4), sterile-filtered, and stored at −80°C until further use. For NMR studies, the SEC buffer was replaced with 100 mM NaCl, 50 mM HEPES, pH 7.4. Determination of proteins concentration was done by measuring absorbance at 280 nm, applying an extinction coefficient of 5,960 M^-^¹cm^-^¹.

### Liposome preparation

5 mg of 1,2-Dioleoyl-sn-glycero-3-phosphoethanolamine (DOPE):1,2-dioleoyl-sn-glycero-3- phospho-L-serine (DOPS):1,2-dioleoyl-sn-glycero-3-phosphocholine (DOPC) 5:3:2 w/w (Avanti Polar Lipids) were resuspended in 0.8 mL methanol:chloroform 1:1, evaporated under a nitrogen stream, lyophilized o/n and resuspended in 0.52 mL of HEPES buffer (50 mM HEPES, 100 mM NaCl, pH 7.4, 0.02% NaN_3_). The resulting turbid sample was sonicated until transparency (15 minutes, 10’’ on, 20’’ off).

### Circular dichroism (CD) spectroscopy

Individual WT and K58N αSyn-to-lipid ratios were pipetted (ratio: 1:10, 1:20, 1:40, 1:100 and 1:200) at a protein concentration of 50 µM and the respective lipid concentrations from a 12.5 mM liposome stock in HEPES buffer (50 mM HEPES, 100 mM NaCl, pH 7.4, 0.02% NaN_3_) to a total volume of 12 µL. For CD measurement the sample was diluted in 48 µL deionized water and transferred to a 0.02 cm pathlength FireflySci cuvette for a final protein concentration of 10 µM. CD data were collected from 190 to 260 nm by using a Chirascan-plus qCD spectrometer (Applied Photophysics, Randalls Rd, Leatherhead, UK) at 20 °C, 1.5 time-per-point (s) in 1 nm steps. The datasets were averaged from three repeats. All spectra were baseline corrected against buffer in deionized water and smoothened (window size: 8).

### NMR spectroscopy

NMR experiments were measured on a Bruker 900 MHz spectrometer equipped with a 5 mm triple-resonance, pulsed-field z-gradient cryoprobe. Two-dimensional ^1^H,^15^N and ^1^H,^13^C heteronuclear single quantum coherence (HSQC) and ^1^H,^1^H total correlation spectroscopy (TOCSY) experiments were acquired for monomer characterization at 15 °C. All experiments were performed in HEPES buffer (50 mM HEPES, 100 mM NaCl, pH 7.4, 0.02% NaN_3_) with 5 % (v/v) D_2_O. Spectra were processed with TopSpin 3.6.1 (Bruker) and analyzed using Sparky 3.115 (T. D. Goddard and D. G. Kneller, SPARKY 3, University of California, San Francisco). The combined HN/N chemical shift perturbation was calculated according to (((δHN)^2^+ (δN/10)^2^)/2)^1/2^. Secondary structure was calculated subjecting the experimental HA, CA, HN and N chemical shifts to TALOS-N (*79*).

### Molecular dynamics simulations

Starting structures of the αSyn WT and K58N peptides (residues 51-65) were built in the PyMOL Molecular Graphics System (Version 1.8.4.0, Schrödinger, LLC). Initially, the peptides were equilibrated in a water box with 50,000 steps of energy minimization. To further equilibrate the system, 100 ps each of volume (NVT) and pressure (NPT) equilibration were performed without position restrains. The MD simulations were carried out in GROMACS (version 2018.3) using the AMBER99SB-ILDN force field and the TIP3P water model at a temperature of 300 K, 1 bar of pressure and with a coupling time (ζT) of 0.1 ps. The peptides were solvated in water with 150 mM NaCl, ensuring overall charge neutrality. The particle mesh Ewald algorithm was used for calculation of the electrostatic term, with a radius of 16 Å for the grid-spacing and Fast Fourier Transform. The cut-off algorithm was applied for the non-coulombic potential with a radius of 10 Å. The LINCS algorithm was used to contain bonds and angles. MD simulations were performed during 100 ns in 2 fs steps and saving the coordinates of the system every 10 ps. The α-helical content over the simulation trajectory was analyzed using the PyMOL Molecular Graphics System. Error bars were calculated from the results of 40 peptides (5 peptides in the water box of 8 independent simulations).

### In vitro thioflavin T fluorescence-based aggregation assays

For the ThT aggregation kinetics assay setup, lyophilized protein was reconstituted in sterile filtered bidistilled water. To get rid of any potential aggregates in protein solutions, samples were first centrifuged at 14,000 rpm for 5 minutes in 100 kDa MWCO filter tubes (Sigma-Aldrich, MO, USA) to collect the filtrate containing monomeric aSyn. Protein concentration was determined on an LVis Plate (BMG Labtech; Ortenberg, Germany) using a CLARIOstar Plus plate reader (BMG Labtech; Ortenberg, Germany) employing the previously mentioned extinction coefficient for aSyn. Prior to initiating the assay, a master mix of 0.5 mg/mL WT or K58N aSyn was prepared in 150 mM NaCl, 10 mM PBS (pH 7.4), 1 mM EDTA, 0.002% SDS, and 25 µM ThT, with 100 µL added into each well in quadruplicate per condition, in addition to the use of protein-free master mix as a blank. The aggregation assay was conducted using CLARIOstar Plus plate reader (BMG Labtech, Ortenberg, Germany) and Costar black, clear-bottom 96-well half-area plates with preloading a single 1-mm glass bead to each well. Plates were sealed with microplate tape and transferred to the reader, with the following settings: orbital shaking (60 seconds on, 30 seconds off) at 400 rpm, 37 °C, in 3.66 min cycles for a total of 1,000 cycles. ThT fluorescence was measured at the end of each cycle with bottom optics, excitation at 450 ± 10 nm, and emission at 480 ± 10 nm. Aggregation curves were then blank-corrected and normalized to the maximum fluorescence for each run.

### Cryo-EM imaging and analysis of WT and K58N fibrils

#### Cryo-EM sample Preparation

Copper 200 mesh R2/1 Quantifoil grids were plasma cleaned for 45 seconds at medium power using a Harrick Plasma PDC-32G-2 plasma cleaner and then mounted on a Vitrobot Mark IV (Thermo Fisher). Alpha-synuclein fibrils, prepared at a concentration of 0.5 mg/mL in 30 mM Tris-HCl buffer at pH 7.5, were sonicated in a waterbath sonicator for 10 minutes on medium settings with a 0.5s ON/OFF interval. Shortly thereafter a 3 mL aliquot of the fibrils was applied to the grid. Blotting was performed with a blot force of 7 for 5 seconds. During the freezing process, the Vitrobot chamber was kept at 10°C with 100% humidity.

#### Data Collection

Cryo-EM datasets were acquired using a Titan Krios electron microscope (Thermo Fisher) equipped with a Falcon4i detector operated in counting mode. For this study, we collected a dataset for the K58N aSyn mutant under the described conditions and used a previously acquired WT dataset, originally collected as part of the G14R study, as a control. Both datasets were collected at a nominal magnification of 130,000x, yielding a pixel size of 0.92 Å. The K58N dataset consisted of 5,813 movies, while the WT dataset comprised 4,092 movies. The defocus range for both datasets was maintained between −1.2 and −2.4 µm. A total electron dose of 40 e^-^/Å² was applied, with exposures of 6.59 e^-^/Å²*s for the WT dataset and 6.61 e^-^/Å²*s for the K58N dataset. An energy filter with a 15 eV slit width was employed during data collection.

#### Data Processing

The raw EER movies were fractionated, aligned, and summed using MotionCor2 with a dose per frame of 1 e^-^/Å². Contrast transfer function (CTF) parameters were estimated using CTFFIND4. To select segments for both the WT and K58N datasets, a crYOLO model was employed, which had been trained on previously grown recombinant aSyn filaments. For this, fibrils from about 50 micrographs of a previous WT aSyn dataset were manually picked in RELION-4, and the coordinates were exported to train a picking model in crYOLO. This model was subsequently used to automatically pick fibrils across all micrographs. The filament coordinates were imported into RELION and used to extract helical segments with an inter-box spacing of approximately 15 Å, corresponding to three asymmetric units per segment. Segments were first extracted with a box size of 768 pixels, binned three times to a final box size of 256 pixels, and then subjected to 2D classification. After excluding picking artifacts, such as carbon edges, the remaining particles were used to extract unbinned segments with a box size of 384 pixels and a pixel size of 0.92 Å/px. Another round of 2D classification was performed, and only the classes showing beta-sheets were selected for further analysis. In case of 1 protofilament (PF) classes for the K58N dataset, no further classifications were carried out since class averages did not show betasheets. Segments of the selected classes underwent 3D classifications, during which helical parameters were systematically scanned as the crossover distance was not visible in the micrographs and the twist could not be determined directly. Multiple 3D classifications were carried out with a fixed helical rise of 4.75 Å and a twist ranging from −0.5° to −1.7° in roughly 0.05° increments. A featureless cylinder was used as the initial model. Classes showing distinct polypeptide signals were utilized as initial models for subsequent 3D refinements that were performed using a sampling interval of 1.8° and a T-value of 15 to 30. Postprocessed models were then used for CTF refinements and final 3D refinements to optimize resolution and model quality.

### aSyn aggregation studies in cells

#### Cell culture

Human neuroglioma cells (H4) were maintained in Opti-MEM I Reduced Serum Medium (Life Technologies-Gibco, Carlsbad CA, USA) supplemented with 10% Fetal Bovine Serum Gold (FBS) (PAA, Cölbe, Germany) and 1% Penicillin-Streptomycin (PAN, Aidenbach, Germany). The cells were grown at 37°C in an atmosphere of 5% CO2.

#### Cell Transfection

Twenty-four hours prior to transfection, approximately 80000 H4 cells were plated per well in a 12-well plate (Costar, Corning, New York, USA). Six hours prior to transfection, the medium was replaced with a fresh one. Transfection protocol using Fugene methodology was carried out as described by the manufacturer. Briefly, ratio of 1 (equal amounts of the plasmids encoding SynT Wild Type Kozak or SynT K58N Kozak with/without Synphilin-1-V5) :3 (Fugene solution) mix was prepared in Optimem medium without adds. The mix was incubated for 20 min and added dropwise to the cells while the plate was gently rocked.

#### Immunocytochemistry

Forty hours after transfection, the medium was removed, the cells were washed with PBS and fixed with 4% paraformaldehyde (PFA) for 30 minutes at room temperature (RT). Cell permeabilization with 0.1% Triton X-100 (Sigma-Aldrich, St. Louis, MO, USA) for 20 minutes at RT was performed, followed by blocking in 3% Bovine serum albumin (Nzytech, Lisbon, Portugal) in PBS 1x for 1 hour. Afterwards, cells were incubated with primary antibody mouse anti-ASYN (1:1000, BD Transduction Laboratory, New Jersey, USA) overnight and secondary antibody Alexa Fluor 488 goat anti-mouse (Life Technologies-Invitrogen, Carlsbad, CA, USA) for 2 hours at RT. Finally, cells were stained with DAPI (Carl Roth, Karlsruhe, Germany) (1:5000 in PBS 1x) for 10 minutes, and the coverslips mounted in SuperFrost® Microscope Slides treated with Mowiol (Calbiochem, San Diego, CA) dried and stored at room temperature until further visualization and analysis. Images were acquired using a confocal point-scanning microscope (Zeiss LSM 900 with Airyscan, Carl Zeiss, Jena, Germany). For each condition, 50 images of the 10 slices Z-Stack were taken using the 63x objective (Objective Plan-Apochromat 63x/1.4 Oil DIC M27) and specific definitions for each staining in the ZEN Software (Carl Zeiss, Jena, Germany).To quantify the number of aggregates and the size of the aggregates (area of the inclusions) inside the cells, the aSyn channel was selected and the images were thresholded The images were then analysed using the Analyse particle plugin from Fiji open-source software (*80*).

### Solubility and dynamic pS129 reversibility experiments

Plasmids and lentivirus production: synthetic cDNA sequences encoding WT or K58N aSyn were digested at the SpeI/NotI enzymes restriction sites and then ligated into pLVX-EF1a-IRES ZsGreen1 vector (TaKaRa) for their expression, driven by the EF1a promoter. Lentiviral packaging was carried out in 293-T cells as previously described (*34*). Briefly, 293-T cells were transfected with plasmids encoding WT or K58N aSyn along with the packaging plasmids pMD2.G and psPAX2 (Addgene #12259 and #12260). After transfection, the viral particles from the culture supernatant were collected, and subjected to ultracentrifugation at 100,000g for obtaining purified/concentrated viral particles. The purified viral pellet then reconstituted in neurobasal medium containing B-27 and Glutamax (Gibco), resulting in a yield around 2·5×10^6^ viral particles per μL. To investigate the impact of K58N mutation on phosphorylation status of aSyn at S129, we cultured primary cortical neurons from E18 pregnant SNCA knockout (SNCA−/−) rats on 24-well plates previously coated with poly-d-lysine, and induced the expression of human WT or K58N proteins by lentiviral transduction at DIV5 as described (*49*). To assess the solubility of aSyn of K58N vs. WT aSyn, we performed sequential protein extraction to isolate cytosol (C) vs. membrane (M) protein fractions using the on-plate extraction technique as previously described (*81*). For the experiments assessing dynamic reversibility of pS129, cortical cultures were treated at DIV17-21 with vehicle (DMSO), 20 µM picrotoxin (PTX), 1µM tetrodotoxin (TTX), or combination of both PTX and TTX. Following treatment for 2h or 4h intervals, cell lysis and immunoblotting was conducted to measure the levels of total and pS129 aSyn as previously described (*34*, *49*). For this study, at least three independent experiments were carried out on different days, with a total of 10-16 biological replicates. Data presented in Fig. 6 B, D, and I-L are statistically analyzed by an unpaired *t*-test with Welch’s correction, while data in F-G were analyzed by Brown-Forsythe and Welch ANOVA with Dunnett’s T3 *post hoc* test for multiple comparisons. Data in Fig. 6 H were analyzed by a two-way ANOVA with Šídák’s multiple comparisons test. Please note: In the experiments assessing the solubility and dynamic reversibility of pS129, both G14R (*24*) and G58N mutants were characterized in parallel. Consequently, the data points for WT-aSyn are identical across both sets of experiments (Fig. 6 D, F, H, I, J, K, and L).

### aSyn expression and purification for phase separation studies

Recombinant wild-type and K58N human full-length aSyn was expressed in BL21(DE3) competent Escherichia coli (C2527, NEB, Ipswich, US) using vector pET28a (Addgene #178032). Bacteria were cultured in LB media supplemented with 50 μg/mL kanamycin (37 °C, constant shaking at 250 rpm). Expression was induced at an OD600 of 0.8 using 1mM isopropyl β-D-1-thiogalactopyranoside (IPTG) and cultured overnight at 25 °C. Cell pellets were harvested by centrifugation at 4000 × g for 30 minutes (AVANTI J-26, Beckman Coulter, USA). aSyn was purified using a protocol previously described (2). Briefly, the cell pellet was resuspended in lysis buffer (10 mM Tris, 1 mM EDTA, Roche cOmplete EDTA free protease inhibitor cocktail, pH 8). The cells were disrupted using a cell disruptor (Constant Systems, Daventry, UK) and were ultracentrifuged at 4 °C, 186,000 × g for 20 minutes (Ti-45 rotor, Optima XPN 90, Beckman Coulter, USA). The supernatant was collected and heated for 20 minutes at 70 °C to precipitate heat-sensitive proteins, followed by ultracentrifugation as above. Streptomycin sulfate (5711, EMD Millipore, Darmstadt, Germany) was added at a final concentration of 10 mg/mL to the supernatant and continuously stirred at 4 °C for 15 minutes to precipitate DNA, followed by ultracentrifugation as above. Ammonium sulfate (434380010, Thermo Scientific) was added at a final concentration of 360 mg/mL to the supernatant and continuously stirred at 4 °C for 30 minutes to precipitate the protein. The precipitated protein was then centrifuged at 500 × g for 15 min, dissolved in 25 mM Tris, pH 7.7, and dialyzed overnight against the same buffer to remove salts. The protein was purified using ion exchange on a HiTrapTMQ HP 5mL anion exchange column (17115401, Cytiva, Sweden) using gradient elution with 0-1M NaCl in 25 mM Tris, pH 7.7. The collected protein fractions were run on SDS-PAGE and pooled fractions were further purified using size-exclusion chromatography on a HiLoadTM 16/600 SuperdexTM 75 pg column (28989333, Cytiva, Sweden). The fractions were collected, and their purity was confirmed using SDS-PAGE analysis. Protein concentrations were determined by measuring absorbance at 280 nm using an extinction coefficient of 5,600 M^-1^ cm^-1^. The monomeric protein was frozen in liquid nitrogen and stored in 25 mM HEPES buffer pH 7.4 at −70 °C. pET28a Cdk2ap1CAN was a gift from Lin He (Addgene plasmid # 178032; http://n2t.net/addgene:178032; RRID:Addgene 178032) (3).

### aSyn labelling

Labelling of aSyn was performed in bicarbonate buffer (C3041, Sigma) at pH 8 using NHS-ester active fluorescent dye AlexaFluor 488 5-SDP ester (A30052, Invitrogen Thermo Fisher). Excess-free dye was removed by buffer exchange using PD10 desalting columns (IP-0107-Z050.0-001, emp BIOTECH, Generon). Labelled protein concentrations were estimated using the molar extinction coefficient ε494 nm = 72,000 M^-1^ cm^-1^.

### aSyn phase separation assays

All aSyn phase separation assays were performed in 25 mM HEPES, pH 7.4 unless mentioned otherwise. Phase separation was induced by mixing aSyn and PEG-8000 (BP223, Fisher Bioreagent) in the presence of calcium (21108, Sigma) as indicated. Images for phase-separated samples were acquired on an LSM780 confocal microscope (Zeiss, Oberkochen, Germany) using a 63x oil immersion objective. Zen 2.3 (black edition) and Zen 2.6 (blue edition) were used for data collection and image export. Images were taken at the indicated aSyn concentration, where aSyn was supplemented with 1% Alexa 488 labelled aSyn. For turbidity measurements phase separation samples were set up as described above using indicated concentrations of aSyn and PEG-8000 in the presence of calcium (21108, Sigma). The turbidity of the samples was measured at 350 nm, 25 °C using 96-well Greiner optical bottom plates on a CLARIOstar plate reader (BMG LABTECH, Ortenberg, Germany) under quiescent conditions. CLARIOStar 5.01 was used for data acquisition. A sample volume of 100 µL was used, and readings were taken within 5 minutes of sample preparation. Raw turbidity data are plotted with background subtraction using GraphPad Prism 9.3.1. Data were obtained from four independent repeats.

### Plasmids

Wild-type human full-length SNCA and VAMP2, encoding aSyn and VAMP2, were cloned from cDNA obtained from human neuroblastoma cells (SH-SY5Y) and inserted into the pEYFP-N1 and pMD2.G vector (Addgene #96808, #12259) with a C-terminal YFP and Flag-tag, respectively. aSyn K58N was generated using KLD substitution (M0554S, NEB, Ipswich, US). All sequences were verified by sequencing. 5HT6-YFP-Inpp5e was a gift from T. Inoue (Addgene plasmid 96808; RRID: Addgene_96808)128. pMD2.G was a gift from D.Trono (Addgene plasmid 12259; RRID: Addgene_12259).

### Cell culture and transfection

HeLa cells were obtained from the European Collection of Cell Cultures (ECACC 93021013) and grown in Dulbecco’s modified Eagle’s Medium (DMEM) high glucose (31966-021, Gibco) supplemented with 10% fetal bovine serum (FBS, F7524, Sigma) and 1% Penicillin/Streptomycin (P0781, Sigma). Cells were grown at 37 °C in a humidified incubator with 5% CO2. Cells were tested for mycoplasma contamination using MycoStripTM (IvivoGen, Toulouse, France). Cells were plated at 20,000 cells/well in 8-well ibidi dishes (80807, ibidi, Gräfelfing, Germany) for confocal imaging or in 48-well plates (Cellstar, 677 180, Greiner bio-one) for incuCyte experiments. Cells were transfected the following day using Fugene HD Transfection reagent according to the manufacturer’s protocol (E2311, Promega). Briefly, per reaction 12.5 μL OptiMEM (31985-062, Gibco) were set up in 1.5 mL sterile Eppendorf tubes. A total of 250 ng of DNA and 0.75 μL of Fugene reagent were added and incubated for 15 min at room temperature. The transfection mix was added to the cells for 1 min and then topped up with 300 μL complete media. Cells were imaged the next day.

### Confocal microscopy and IncuCyte

Live cell confocal imaging was performed on an LSM780 microscope (Zeiss, Oberkochen, Germany) using a 63x oil immersion objective. YFP fluorescence was excited with the 514 laser at 2% laser power. Zen 2.3 (black edition) and Zen 2.6 (blue edition) were used for data collection and image export. For quantitative evaluation of condensate formation cells were imaged with the IncuCyte S3 (Essen BioScience, Newark, UK). Phase brightfield and green fluorescence images were taken using a 20x objective at a 4-hour interval at 200 ms exposure, condensate formation (% of cells showing condensate formation) was evaluated 16 hours after transfection. IncuCyte 2021A was used for data analysis. At least three biological repeats with three technical repeats each were analysed blinded to the investigator.

### Quantification and statistical analysis

Data analysis and statistical analysis was performed using Excel 2016 and GraphPad Prism 9.3.1. All data are represented as mean ± standard error (SD) if not indicated otherwise. Statistical analysis was carried out using unpaired two-tailed t-test. Statistical parameters are reported in the Fig.s and the corresponding Fig. Legends. Exact p-values are shown. Data distribution was assumed to be normal but this was not formally tested. No statistical methods were used to pre-determine sample sizes but our sample sizes are similar to those reported in previous publications (4–6). Samples were randomly allocated into experimental groups. Data collection and analysis have been performed blinded when indicated. Data were included if the control (wild-type) showed appropriate condensate formation.

### Yeast Plasmids

The aSyn-K58N variant was constructed by site-directed mutagenesis using the QuickChange II Site-Directed Mutagenesis Kit (Agilent Technologies, SC, USA), in the plasmid backbone p426GPD encoding the WT aSyn-GFP, following the manufacturer’s instructions. We also constructed a plasmid expressing only GFP as a control, by introducing the GFP coding sequence as a SpeI-XhoI digested PCR product in the p426GPD backbone. All constructs were confirmed by DNA sequencing.

### Yeast cell growth conditions, viability assay and fluorescence microscopy

The Saccharomyces cerevisiae yeast strain BY4741 (MATa his3Δ1 leu2Δ0 met15Δ0 ura3Δ0) was transformed with GFP, WT aSyn and K58N plasmids by standard lithium acetate method. All strains were grown overnight at 30 °C 180 rpm in yeast minimal synthetic defined (SD) medium (Takara Bio), supplemented with a drop-out mix (Takara Bio) lacking the amino acid uracil (SD- URA) for transformant selection, at a volume/medium ratio of 5:1.

The assessment of cellular viability was achieved by spotting assay. Here, cultures grown to mid-log phase were standardized to equal cellular densities, serially diluted 10-fold starting with an OD600nm of 1 and spotted on SD-URA agar plates. Following 3 days incubation at 30 °C the plates were photographed.

aSyn cellular localization was evaluated by fluorescent microscopy. Images were attained with an epifluorescence microscope Zeiss Axio Observer equipped with a 100x oil objective lens.

## Data Availability

All data produced in the present work are contained in the manuscript. The micrographs used for the single-particle analysis (SPA) of alpha-synuclein fibrils are available in the EMPIAR database under accession code EMPIAR-12518.
The atomic model and cryo-EM density map for the wild-type (WT) alpha-synuclein fibril are deposited in the Protein Data Bank (PDB) and Electron Microscopy Data Bank (EMDB) under accession codes 9HGS and EMD-52165, respectively. The corresponding data for the K58N mutant alpha-synuclein fibril are available under accession codes 9HXA (PDB) and EMD-52458 (EMDB).
A comprehensive Key Resource Table, detailing datasets, software, and protocols is available via Zenodo at https://zenodo.org/records/14730688. Additionally, the entry includes an Excel file with tabular data on the protofilament distribution of WT and K58N alpha-synuclein filaments, XML files containing tabular data for the FSC graphs of each density map used to determine the final resolution (generated with RELION 4.0), and two Python scripts for graphing the FSC XML data.
The full single-particle analysis protocol describing the cryo-EM data processing strategy is available at Protocols.io (https://www.protocols.io/view/single-particle-analysis-of-synuclein-fibrils-81wgbxozylpk/v1).

https://www.protocols.io/view/single-particle-analysis-of-synuclein-fibrils-81wgbxozylpk/v1

## Abbreviations

aSyn: Alpha-synuclein
EOPD: Early onset PD
LBD: Lewy body dementia
MSA: Multiple system atrophy
PD: Parkinson’s disease
*SNCA*: Alpha-synuclein gene

## Acknowledgements

We thank Dr. Ellen Gerhardt for assistance with site-directed mutagenesis in the aSyn-encoding plasmids.

## Author contributions

TFO and BM contributed to the study conception and design. MAA, SB, NR, MR, LS, GM, MZ, KS, AIO, VJ, LA, AA, and AC contributed to the acquisition of data.

MAA, SB, NR, KS, UF, JL, MZZ, CF, JW, CT, MS, SP, RFB, BM, and TFO analyzed and interpreted the data.

MAA, SB, and TFO wrote the initial draft of the manuscript. All authors reviewed and revised the manuscript.

## Funding

T.F.O. is supported by the Deutsche Forschungsgemeinschaft (DFG, German Research Foundation) under Germany’s Excellence Strategy, EXC 2067/1- 390729940, and by SFB1286 (B8). J.L. is supported by the Royal Society (Royal Society Dorothy Hodgkin Research Fellowship, DHF/R1/201228), the Addenbrooke’s Charitable Trust (Grant Award 900325), the Leverhulme Trust (Research Project Grant, RPG-2022-257), as well as a Career Support Fund from the University of Cambridge. L.S. was supported by the Foundation for Science and Technology (FCT), SFRH/BD/143286/2019. N.R. and U.D. are supported by the National Institutes of Health (grant numbers NS121826, NS099328, NS109209, NS122880, and NS133979). M. Zweckstetter is supported by the Michael J. Fox foundation through MJFF- 022411. M.Zech acknowledges grant support by the European Joint Programme on Rare Diseases (EJP RD Joint Transnational Call 2022), and the German Federal Ministry of Education and Research (BMBF, Bonn, Germany), awarded to the project PreDYT (PREdictive biomarkers in DYsTonia, 01GM2302), by the Federal Ministry of Education and Research (BMBF) and the Free State of Bavaria under the Excellence Strategy of the Federal Government and the Länder, as well as by the Technical University of Munich – Institute for Advanced Study. MZ’s research is supported by a “Schlüsselprojekt” grant from the Else Kröner-Fresenius-Stiftung (2022_EKSE.185).

R.F.B. is supported by the Deutsche Forschungsgemeinschaft (DFG, German Research Foundation) under Germany’s Excellence Strategy, EXC 2067/1- 390729940, and by SFB1286 (A12). Cryo-EM instrumentation at the University of Göttingen was jointly funded by the DFG Major Research Instrumentation program (448415290) and the Ministry of Science and Culture of the State of Lower Saxony. K.S. and R.F.-B. were funded by the joint efforts of The Michael J. Fox Foundation for Parkinson’s Research (MJFF) and the Aligning Science Across Parkinson’s (ASAP) initiative. MJFF administers the grant ASAP-000282 on behalf of ASAP and itself.

## Conflict of interests

The authors declare that they have no competing interests to disclose related to the article.

## Data availability

All data needed to evaluate the conclusions in the paper are present in the paper and/or the Supplementary Materials.

The micrographs used for the single-particle analysis (SPA) of alpha-synuclein fibrils are available in the EMPIAR database under accession code EMPIAR-12518.

The atomic model and cryo-EM density map for the wild-type (WT) alpha-synuclein fibril are deposited in the Protein Data Bank (PDB) and Electron Microscopy Data Bank (EMDB) under accession codes 9HGS and EMD-52165, respectively. The corresponding data for the K58N mutant alpha-synuclein fibril are available under accession codes 9HXA (PDB) and EMD-52458 (EMDB).

A comprehensive Key Resource Table, detailing datasets, software, and protocols is available via Zenodo at https://zenodo.org/records/14730688. Additionally, the entry includes an Excel file with tabular data on the protofilament distribution of WT and K58N alpha-synuclein filaments, XML files containing tabular data for the FSC graphs of each density map used to determine the final resolution (generated with RELION 4.0), and two Python scripts for graphing the FSC XML data.

The full single-particle analysis protocol describing the cryo-EM data processing strategy is available at Protocols.io (https://www.protocols.io/view/single-particle-analysis-of-synuclein-fibrils-81wgbxozylpk/v1).

## Notes

### Competing Interest Statement

The authors have declared no competing interest.

### Author Declarations

Written informed consent was given by the patient to participate in a genetic study in cooperation with the Institute of Human Genetics, Technical University Munich, Germany. The study was approved by the Ethics Commission of the Landesärztekammer Hessen, Frankfurt am Main, Germany. Nr: MC 284/2014. Additional consent to confirm the genetic finding at the Institute of Human Genetics, University Clinic Göttingen, Germany was provided.

### Summary of Updates

the manuscript is missing Figure 7. Therefore, the update is just to add Figure 7.

