## Supplementary Material for "A novel alpha-synuclein K58N missense variant in a patient with Parkinson’s disease"

### Supplementary Materials

**This PDF file includes:**

Figs. S1 to S6

**Fig. S1.**

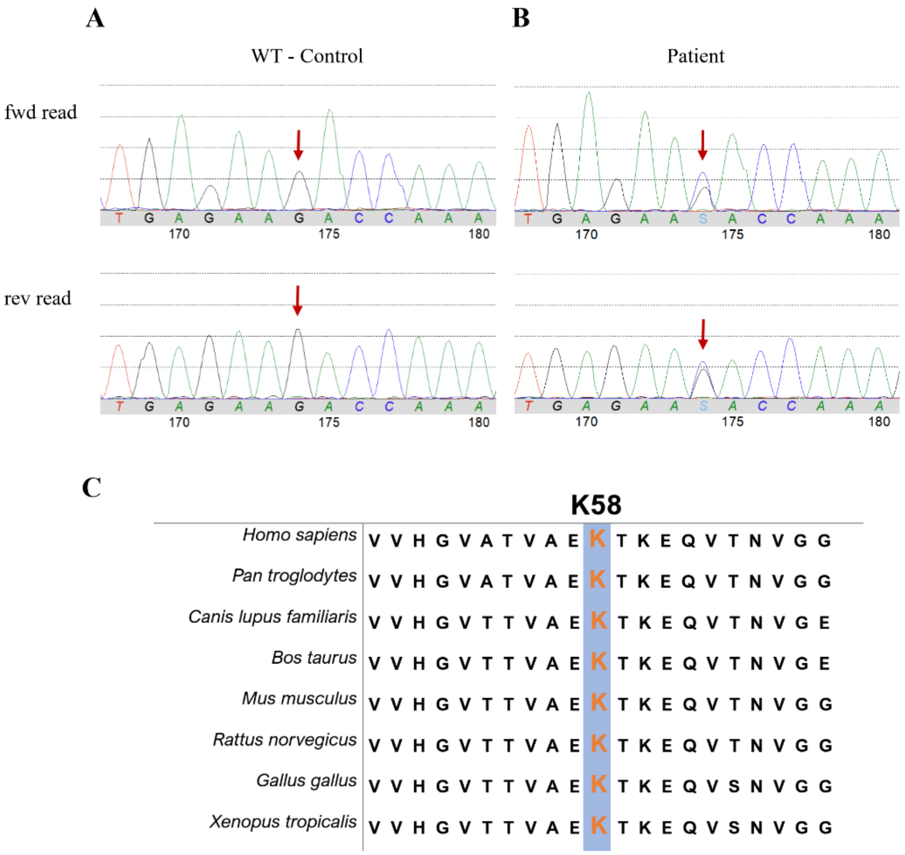


**Fig. S1. Identification of K58N mutation**. (A) and (B) present DNA sequencing results of the SNCA gene, Exon 4, from a wildtype control sample (A) and a patient sample (B) with a heterozygous variant c.174G>C; S = G or C. (C) illustrates conservation of the SNCA K58N missense mutation across various species.

**Fig. S2.**

A

| Parameters | WT aSyn | K58N aSyn |
|---|---|---|
| Length | 140 | 140 |
| #Amyloids | 20 | 20 |
| Best energy | -7.239327 | -7.239327 |
| % disorder | 34.28 | 34.28 |
| % α-helix | 26.43 | 20.71 |
| % β-strand | 22.14 | 25.71 |
| %coil | 51.43 | 53.57 |

B

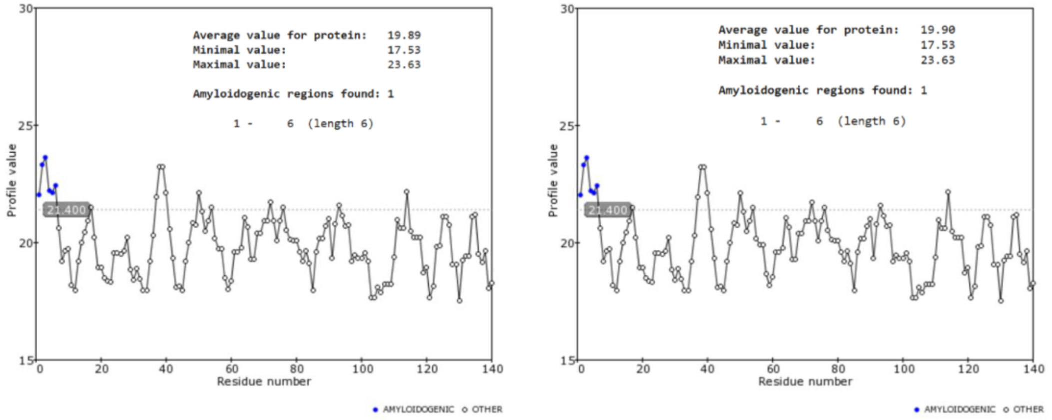


C

| No. | Peptide | Amyloid probability | Non-amyloid probability |
|---|---|---|---|
| 53 | ATVAEK | 0.740 | 0.260 |
| 54 | TVAEKT | 0.172 | 0.828 |
| 55 | VAEKTK | 0.971 | 0.029 |
| 56 | AEKTKE | 0.823 | 0.177 |
| 57 | EKTKEQ | 0.989 | 0.011 |
| 58 | KTKEQV | 0.999 | 0.001 |

D

| No. | Peptide | Amyloid probability | Non-amyloid probability |
|---|---|---|---|
| 53 | ATVAEN | 0.992 | 0.008 |
| 54 | TVAENT | 0.771 | 0.229 |
| 55 | VAENTK | 0.954 | 0.046 |
| 56 | AENTKE | 0.024 | 0.976 |
| 57 | ENTKEQ | 0.999 | 0.001 |
| 58 | NTKEQV | 1.000 | 0.000 |

**Fig. S2. Prediction of the impact of the K58N mutation on structural and aggregation properties of aSyn.** (A) evaluation of the effect of K58N using PASTA 2.0 algorithm. The mutation tends to reduce the amount of α-helix and number coils, and to increase the β-strand content. (B) Prediction of amyloidogenic sequences of WT (left) and K58N (right) aSyn through FoldAmyloid tool based on the probability of formation of hydrogen bonds. A slight increase in the aggregation probability was observed for K58N variant. (C) Analysis of aggregation-prone regions for WT and and K58N aSyn (D) utilizing GAP (Aggregation Proneness) prediction algorithm, which is based on analyzing each part of the protein as a 6-amino acid long peptide and indicating which is the probability of the peptide forming an amyloid structure. According to GAP, there is an increase in the individual probability of each peptide to form amyloid structures, with exception to the peptide in the position 56, for K58N compared to WT aSyn.

**Fig. S3.**

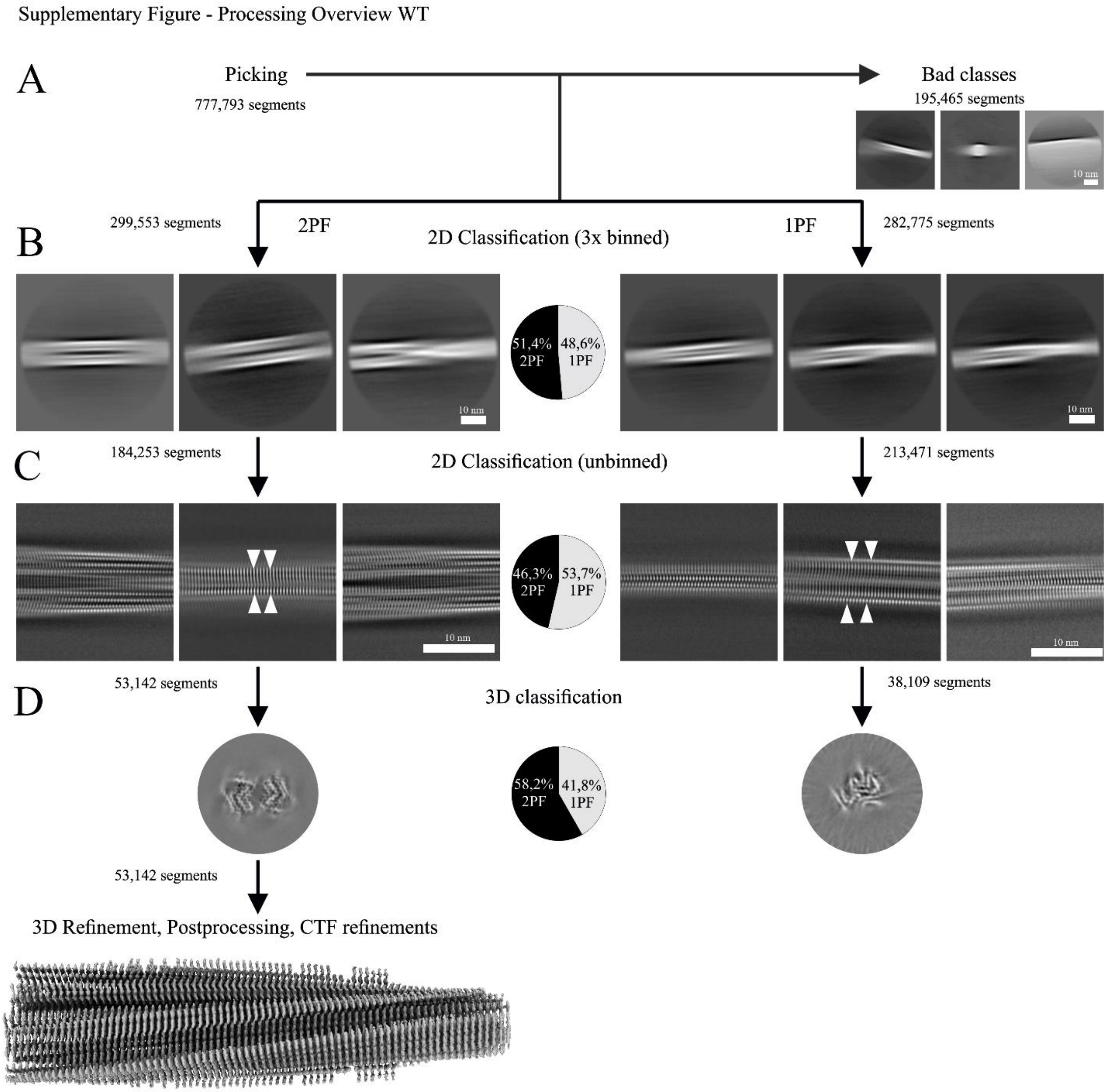


**Fig. S3. Detailed Processing Workflow for the WT aSyn Dataset.** (A) From the initially picked helical segments, a subset was excluded due to artifacts, such as the presence of carbon edges. The remaining segments were sorted into two categories based on structural features: segments exhibiting two protofilaments (2PF, "wide") and those with a single protofilament (1PF, "narrow"). (B) During the initial classification step, which utilized thrice-binned segments, the distribution of segments between the two groups was approximately equal, as depicted in the associated pie chart. (C) Further classification performed on unbinned data confirmed the results of the initial classification, with segments evenly divided between the 2PF and 1PF categories, as illustrated in the corresponding pie chart. (D) The three-dimensional classification results are shown, including the final electron density map of the 2PF data following 3D refinement, postprocessing, and CTF refinement. The accompanying pie chart reveals a slightly greater contribution of segments to the 2PF (two-protofilament) structure compared to the 1PF structure.

**Fig. S4.**

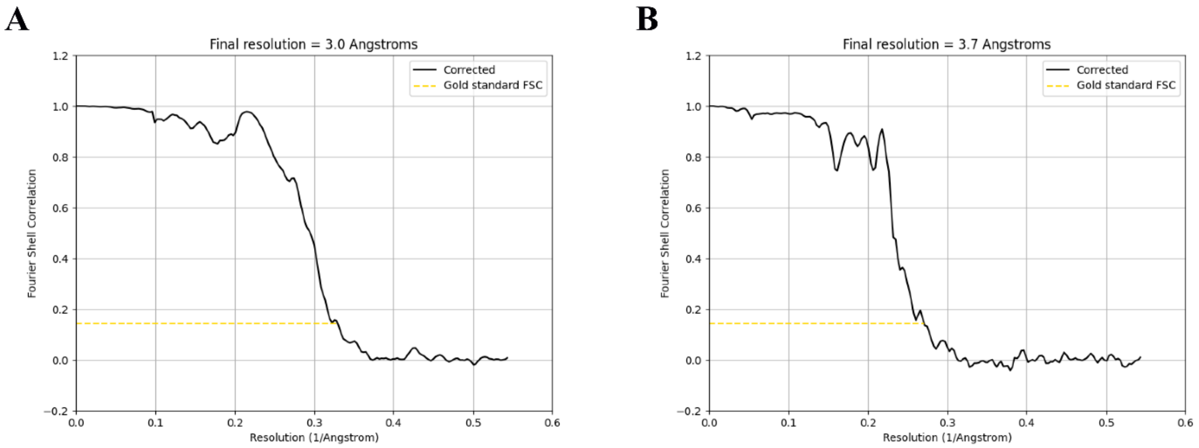


**Fig. S4. Fourier Shell Correlation (FSC) curves for 2PF WT (A) and 2PF K58N (B).**

**Fig. S5.**

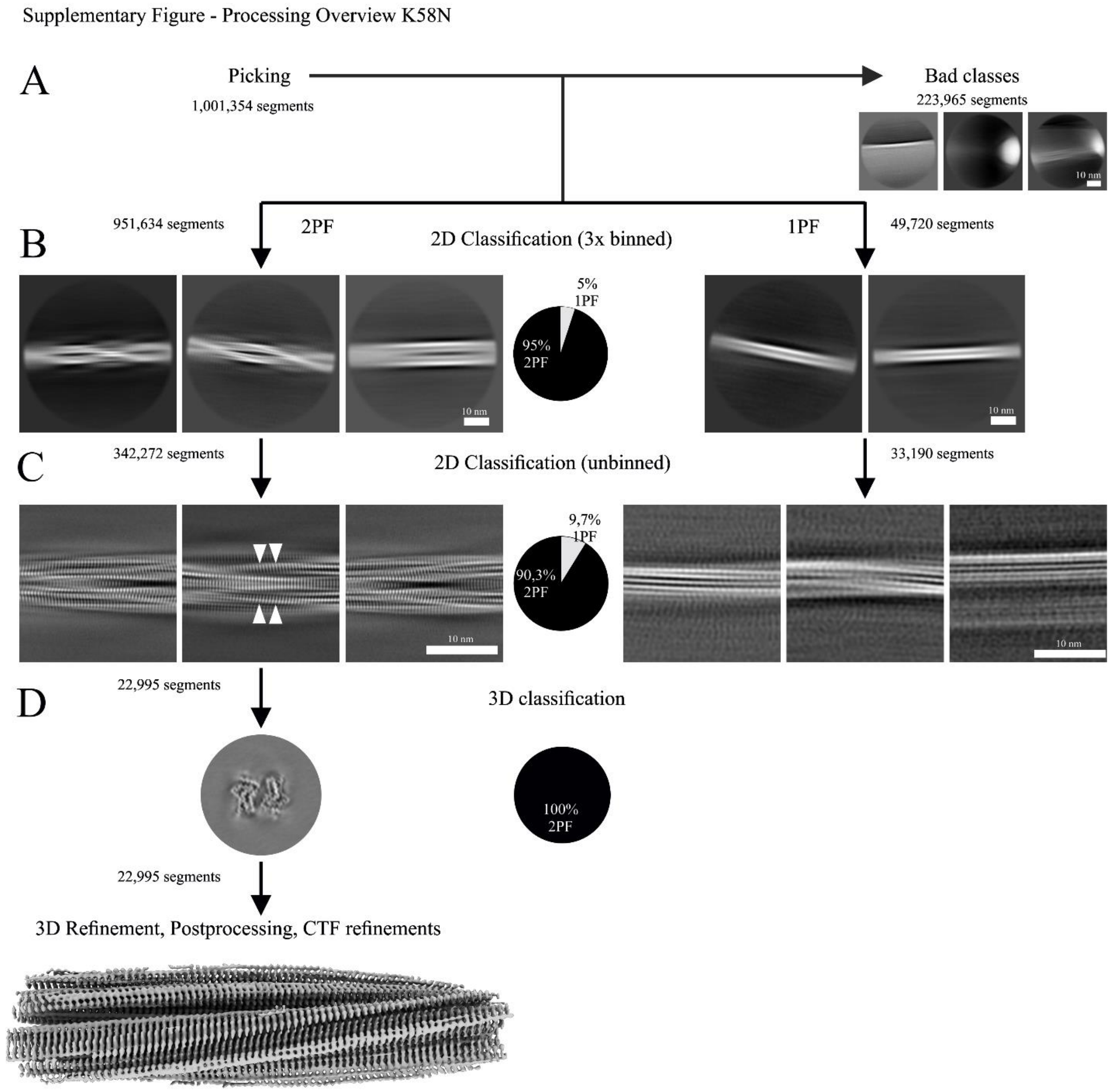


**Fig. S5. Detailed Processing Workflow for the K58N aSyn Dataset.** (A) A subset of the initially picked helical segments was removed due to artifacts, such as those containing carbon edges. The remaining segments were divided into two categories based on their structural features: segments showing two protofilaments (2PF, "wide") and those showing a single protofilament (1PF, "narrow"). (B) During the initial classification step with three times binned segments, a significant majority of the segments were categorized as 2PF, with only a small fraction classified as 1PF, as illustrated by the provided pie chart. (C) Subsequent classification with unbinned data supported the initial results, as depicted by the adjacent pie chart. In contrast ot the 2PF dataset, no clear beta-sheet separation was observed in the unbinned class averages for the 1PF dataset, hinting at the lower quality and quantity of the 1PF segments. (D) The results of the three-dimensional classification are presented. For the 2PF data, the refined electron density map is shown after 3D refinement, postprocessing, and CTF refinement. Due to the poor quality of particles and class averages in the 1PF dataset, further processing via 3D classification was not feasible.

**Fig. S6.**

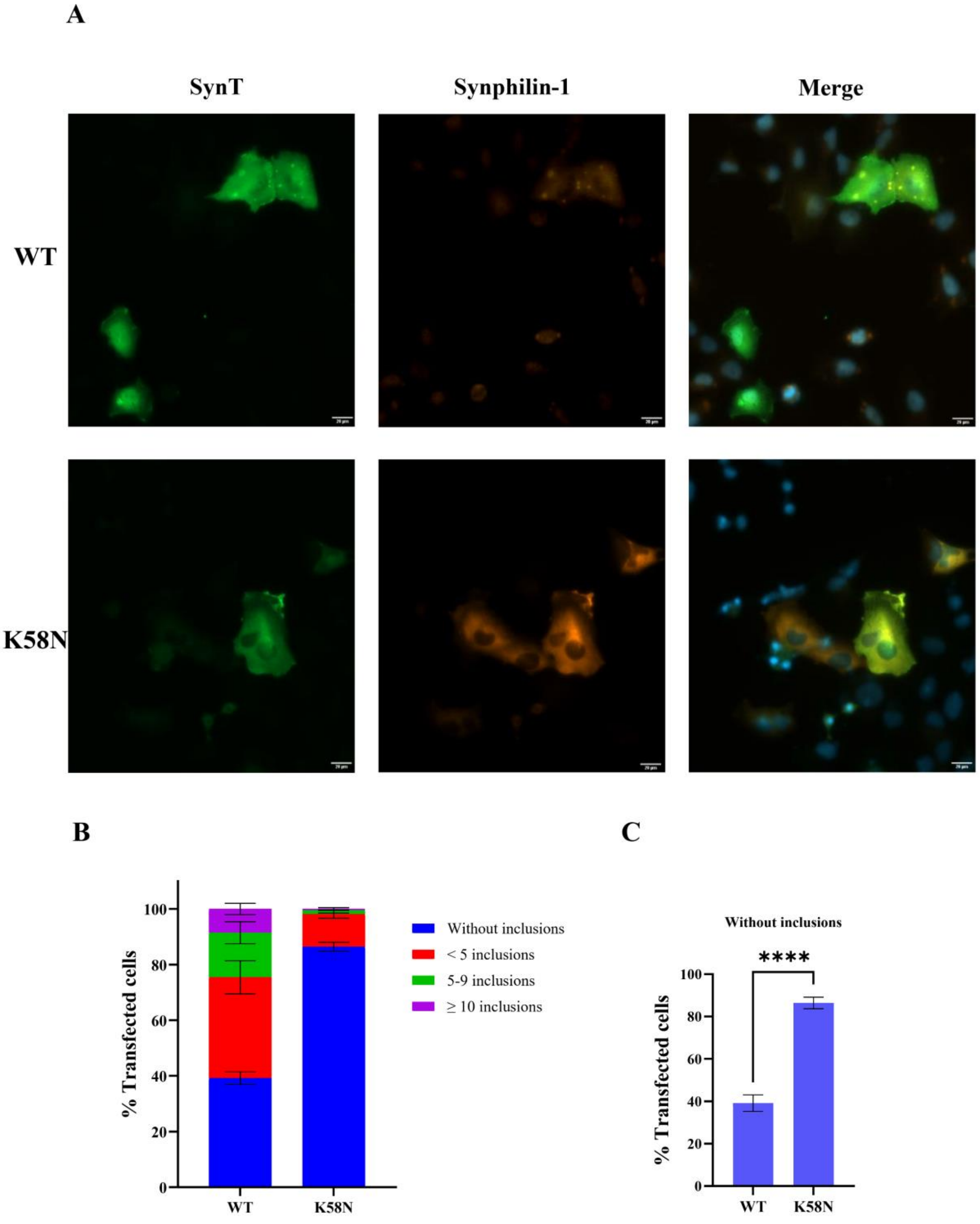


**Fig. S6. Effect of K58N mutation on inclusion formation in cells. (A)** Representative immunohistochemistry images of H4 cells showing the patterns of inclusion formation for both WT and K58N SynT variants. Scale bar: 20 μm. (B) and (C) show the quantification of the number of inclusions from transfected cells. A total of 50 cells were counted using a 20x objective for each experiment, and classified into four groups based on their inclusion patterns. Student's t-test was used to analyze the data, and the results are shown as mean ± SD from 3 independent experiments.
